# Body mass index and risk of COVID-19 diagnosis, hospitalisation, and death: a population-based multi-state cohort analysis including 2,524,926 people in Catalonia, Spain

**DOI:** 10.1101/2020.11.25.20237776

**Authors:** Martina Recalde, Andrea Pistillo, Sergio Fernandez-Bertolin, Elena Roel, Maria Aragon, Heinz Freisling, Daniel Prieto-Alhambra, Edward Burn, Talita Duarte-Salles

## Abstract

**Objective:** To investigate associations between body mass index (BMI) and risk of COVID-19 diagnosis, hospitalisation with COVID-19, and COVID-19-related death, accounting for potential effect modification by age and sex.

**Design:** Population-based cohort study.

**Setting:** Primary care records covering >80% of the Catalonian population (Spain), linked to region-wide testing, hospital, and mortality records from March to May 2020.

**Participants:** People aged ≥18 years with at least one measurement of weight and height from the general population and with at least one year of prior medical history available.

**Main outcome measures:** Cause-specific hazard ratios (HR) with 95% confidence intervals for each outcome.

**Results:** Overall, 2,524,926 participants were followed up for a median of 67 days. A total of 57,443 individuals were diagnosed with COVID-19, 10,862 were hospitalised with COVID-19, and 2,467 had a COVID-19-related death. BMI was positively associated with being diagnosed as well as hospitalised with COVID-19. Compared to a BMI of 22kg/m^2^, the HR (95%CI) of a BMI of 31kg/m^2^ was 1.22 (1.19-1.24) for COVID-19 diagnosis, and 1.88 (1.75-2.03) and 2.01 (1.86-2.18) for hospitalisation without and with a prior outpatient diagnosis, respectively. The relation between BMI and risk of COVID-19 related death was J-shaped. There was a modestly higher risk of death among individuals with BMIs≤19 kg/m^2^ and a more pronounced increasing risk for BMIs ≥37 kg/m^2^ and ≥40 kg/m^2^ among those who were previously hospitalised with COVID-19 and diagnosed with COVID-19 in outpatient settings, respectively. The increase in risk for COVID-19 outcomes was particularly pronounced among younger patients.

**Conclusions:** There is a monotonic association between BMI and COVID-19 infection and hospitalisation risks, but a J-shaped one with mortality. More research is needed to unravel the mechanisms underlying these relationships.

**Summary boxes:** *Section 1: What is already known on this topic:*

- A high body mass index (BMI) has previously been associated in a linear and non-linear fashion with an increased risk of multiple health outcomes; these associations may vary by individual factors such as age and sex.
- Obesity has been identified as a risk factor for COVID-19 severity and mortality. However, the role of general adiposity in relation to COVID-19 outcomes has mostly been studied by dichotomizing BMI (below or above 30 kg/m^2^) or by a diagnostic code indicating obesity.
- Two studies have investigated BMI (as a continuous variable) in relation to COVID-19 outcomes, accounting for non-linearity: one conducted in a tested population sample of the UK Biobank found BMI is related in a dose-response manner with the risk of testing positive for COVID-19; another conducted in a hospital setting in New York reported a J-shaped association between BMI and the risk of intubation or death. These studies were limited in sample size and were prone to collider bias due to the participant’s restriction to tested and hospitalised patients. No studies have described the association between BMI and COVID-19 outcomes across the natural history of the disease (from no disease to symptomatic disease, hospitalisation, and mortality) using data from diverse health settings.

*Section 2: What this study adds:*

- We provide a comprehensive analysis of the association between BMI and the course of the COVID-19 disease in the general population of a Spanish region during the first wave of the pandemic, using linked data capturing outpatient clinical diagnoses, RT-PCR test results, hospitalisations, and mortality (inside and outside of the hospital setting).
- We found that BMI is positively associated with being diagnosed as well as hospitalised with COVID-19, and is linked in a J-shaped fashion with the risk of COVID-19 related death.
- The association between BMI and COVID-19 related outcomes is modified by age and sex; particularly, the risk of COVID-19 outcomes related to increased BMI is higher for those aged between 18 and 59 years, compared to those in older age groups.

## Introduction

The coronavirus disease 2019 (COVID-19), the illness caused by the Severe Acute Respiratory Syndrome Coronavirus 2 (SARS-CoV-2), was declared a global pandemic in March 2020 and has since overwhelmed health care systems worldwide.[1] COVID-19 clinical manifestations range from asymptomatic or mild symptoms to severe illness requiring hospitalisation. A high body mass index (BMI) has previously been associated in a linear and non-linear fashion with an increased risk of multiple health outcomes such as metabolic and cardiovascular conditions, cancer, viral infections, and mortality.[2–5] A better understanding of the relation between BMI and the progression of COVID-19 is essential for clinical management of patients and implementation of preventive strategies.

A review and meta-analysis of 75 studies indicated obesity (defined as a BMI≥30 kg/m^2^) as a risk factor for severe COVID-19 and related mortality.[6] In addition, two studies with data from a subsample of the UK Biobank and a New York hospital found that BMI was associated in a dose-response manner with an increased risk of testing positive and in a J-shaped fashion with the risk of intubation or death, respectively.[7,8] Although these studies have provided relevant insights on this association, they have important limitations that include being restricted to tested or hospitalised populations (increasing the risk of collider bias), having a small sample size, limitedly accounting for potential confounding, or dichotomizing BMI (with vs without obesity).[9] These limitations prevent the generalization of the studies’ conclusions to populations with milder forms of disease or to the general population. A study conducted with comprehensive patient-level data that contains detailed individuals’ BMI information and captures incident COVID-19 cases from a large and representative population, with subsequent longitudinal follow-up, and where outcomes are recorded in diverse healthcare settings, could address the limitations of the previous evidence.

Catalonia was heavily hit by the first phase (March 1st through the first week of May) of the COVID-19 pandemic.[10] This region has a universal taxpayer-funded primary care-based health system in which general practitioners have been the first point of contact for care throughout the pandemic. Electronic health records (EHRs) from primary care encompassing demographic, historical lifestyle information and disease diagnoses linked to SARS-CoV-2 Reverse Transcription Polymerase Chain Reaction (RT-PCR) test results, hospital records, and regional mortality data offer a unique opportunity to study the role of BMI in the course of COVID-19. In this study, we investigated the associations between BMI and risks of COVID-19 diagnosis, hospitalisation with COVID-19, and COVID-19 related death, accounting for potential effect modification by age and sex, using EHR data from Catalonia.

## Methods

### Study design, setting and data sources

We conducted a cohort study from the 1st March 2020 to the 6th May 2020. We used prospectively collected primary care records from the Information System for Research in Primary Care (SIDIAP; www.sidiap.org) in Catalonia, Spain. SIDIAP contains anonymized EHRs for approximately six million people (80% of the Catalan population) since 2006 and is representative of the Catalan population in terms of age, sex, and geographic distribution.[11] SIDIAP includes high-quality data on anthropometric measurements, disease diagnoses, prescription and dispensation of drugs, laboratory tests, demographic and lifestyle information. The SIDIAP database has been linked to COVID-19 RT-PCR test results, hospital records, and regional mortality data, and mapped to the Observational Medical Outcomes Partnership (OMOP) Common Data Model (CDM).[12] The latter allowed to structure the data in a standardised format, and to apply analytical tools developed by the open-science Observational Health Data Sciences and Informatics (OHDSI) network.[13]

### Multistate framework

We addressed our objectives using a multi-state framework. Multi-state models allow for a description of the progression from a time origin until the occurrence of several events, extending on competing risk models by also describing transitions to intermediate events.[14] In the context of COVID-19, outpatient diagnoses of the disease and hospitalisations with the disease can be considered as intermediate events between not being (identified as) infected on one end to death on the other. Therefore, we structured our multi-state model in four states: *general population, diagnosed* (with COVID-19), *hospitalised* (with COVID-19), and *death* (Figure 1). The following transitions were possible: *general population* to either *diagnosed, hospitalised* or *death*; *diagnosed* to either *hospitalised* or *death*; *hospitalised* to *death*.

**Figure 1.**
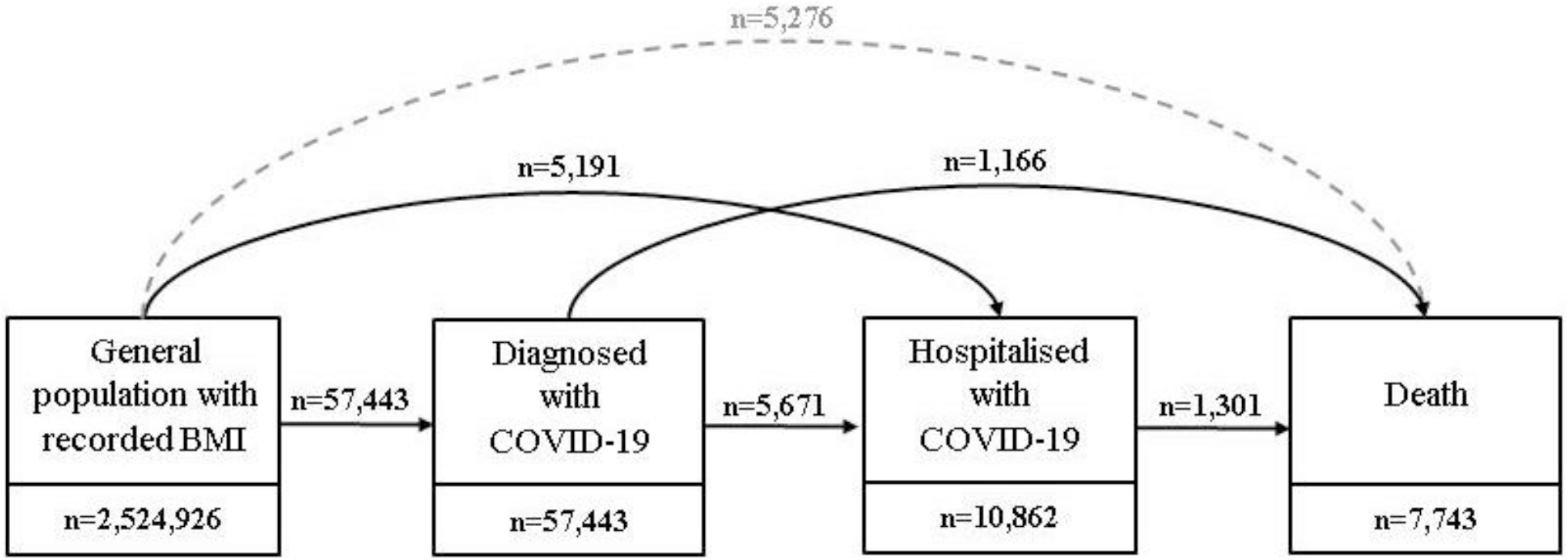
Overview of the multi-state model which provided the framework for this study. Notes: The transition from general population to death was used for censoring, but was not a transition of interest in modelling (grey dashed lines). Abbreviations: BMI: Body Mass Index; COVID-19: Coronavirus Disease 2019.

### Participants

For the primary analyses, we identified all adults (aged 18 years or older) registered in the SIDIAP as of the 1st March 2020 with a BMI recorded at an age equal or greater than 18 years. We included individuals with at least one year of prior history available (because we needed sufficient time to capture participants’ characteristics prior to study entry), without a previous clinical diagnosis or positive test result for COVID-19, who were not hospitalised or living in a nursing home on the 1st March 2020 (to have study participants representative of the community population) and who had information on both smoking and socioeconomic status. The flow chart of inclusion and exclusion criteria for this study is presented in Figure S1. Individuals’ follow-up period began on the 1st of March 2020 (index date) and ended for any given transition due to exit from the database (administrative censoring), the occurrence of the event of interest or a competing event, or the end of the study period (the 6th May 2020).

### Variables

The exposure of interest was BMI as a continuous variable (in kg/m^2^). BMI was calculated using the weight (kg) and height (cm) of patients assessed in a standardized manner by general practitioners or nurses.[15] The exposure was assigned as the closest valid BMI (≥15kg/m^2^ and ≤60kg/m^2^) to the index date recorded between January 1st 2006 and February 29th 2020.

The characteristics of interest were sex, age, smoking status, socioeconomic status, and comorbidities. We extracted participants’ sex (female, male), age (in years) at index date and smoking status (never, former or current smoker). We assessed socioeconomic status using the “Mortalidad en áreas pequeñas Españolas y Desigualdades Socioeconómicas y Ambientales” (MEDEA) deprivation index, which is calculated at the census tract level in urban areas of Catalonia.[16] This measure is categorized into quintiles for anonymization purposes where the first quintile represents the least deprived group of the population and the fifth the most deprived one. It also includes a rural category since the MEDEA index is not available for participants living in those areas. We identified the following comorbidities using the individual’s medical history: autoimmune condition, chronic kidney disease, chronic obstructive pulmonary disease (COPD), heart disease, hyperlipidemia, hypertension, malignant neoplasm (excluding non-melanoma skin cancer) and type 2 diabetes. We selected these conditions based on their relevance to the obesity and COVID-19 research fields and their availability in the OMOP-CDM mapped version of the SIDIAP database.[17,18] We defined these comorbidities as in a previous COVID-19 study conducted using SIDIAP data.[19] The definitions can be consulted in a web application (“Index Event Breakdown” tab) available at https://livedataoxford.shinyapps.io/MultiStateCovidCohorts/.

The outcomes of interest were an outpatient (primary care) clinical diagnosis of COVID-19, a hospitalisation with COVID-19, and death. We defined outpatient COVID-19 diagnoses based on a recorded clinical code for COVID-19 disease (International Classification of Diseases, Tenth Revision, Clinical Modification [ICD-10-CM] codes B34.2 “Coronavirus infection, unspecified” and B97.29 “Other coronavirus as the cause of diseases classified elsewhere”). We did not require a positive RT-PCR test result in the definition of clinical diagnoses of COVID-19 due to testing restrictions during the first months of the pandemic.[19] We defined hospitalisation with COVID-19 as a hospital admission (hospital stay of at least one night) where the individual had a positive RT-PCR test result or a clinical diagnosis of COVID-19 over the 21 days prior to their admission up to the end of their hospital stay. We defined mortality using region-wide mortality data, and so included both deaths during hospitalisations and in the community.

### Statistical analyses

We reported the participants’ baseline characteristics by World Health Organization (WHO) categories of BMI (underweight or normal weight [BMI <18.5 kg/m^2^ and between ≥18.5 and <25 kg/m^2^], overweight [BMI ≥25 and <30 kg/m^2^] and obesity [BMI ≥30 kg/m^2^]).

We compared the baseline characteristics of the included individuals to those of the excluded due to unavailability of BMI, smoking status or the MEDEA deprivation index information using standardized mean differences (SMDs). We considered SMDs >|0.1| indicate meaningful differences in the distribution of a given characteristic between the two groups.[20]

We described the participants’ time at risk at each state and the absolute number of outcomes observed for each transition, by WHO categories of BMI. We assessed the relationship between BMI and the risk of transitioning to a subsequent state in the multistate model by estimating cause-specific hazard ratios (HRs) and 95% confidence intervals (CIs) using Cox proportional hazard regressions. We estimated three types of models: 1) with BMI as the sole explanatory variable (unadjusted models); 2) adjusted for age and sex; 3) adjusted for age, sex, smoking status and the MEDEA deprivation index (fully adjusted models). We used a directed acyclic graph to guide decisions on the control for confounding (Figure S2).[21] We considered non-linearity in BMI and transitions by fitting models with BMI as a linear term, with a polynomial of degree 2 (i.e. quadratic) and with restricted cubic splines (with 3, 4, or 5 knots).[22] We calculated the Bayesian Information Criterion (BIC) and we favoured the model with the lowest BIC values. We compared the model where BMI was fitted with a non-linear term against a linear model using a likelihood ratio test. We fitted age in the adjusted models using the same strategy as we did for BMI. We checked the proportional hazard assumptions for the variables included in the models by visual inspection of log-log survival curves. We did not model the transition from the *general population* to *death* because we were interested in COVID-19-related deaths which we captured by having gone through the *diagnosed* or *hospitalised* states (Figure 1). However, we considered *death* among the *general population* as a competing risk by censoring people at their death.

We assessed effect modification by introducing interaction terms (one at a time) between BMI and age and sex. We stratified the models in three categories of age (18 to 59 years, 60 and 79, and 80 or above) and sex (female and male). As a secondary analysis, we re-estimated the models fitting BMI in WHO categories.

For the main analyses, we conducted a complete case analysis (where we only included individuals with complete information on BMI and the covariates of interest). To explore the possibility of selection bias due to excluding those with missing data, in a sensitivity analysis we re-estimated the main models after multiple imputation (using predictive mean matching, with 5 imputations drawn) of missing data on BMI, smoking status, and the MEDEA deprivation index. In a second sensitivity analysis, we considered the impact of exposure misclassification by replicating the main analyses including only BMI values recorded in the previous five years (from March 1st 2015 to February 29th 2020).

We used R version 3.6 for data analysis and visualization. The R packages used for the analyses included numerous tidyverse packages, mstate, survival and rms.[23–26] The analytic code we used is available at https://github.com/SIDIAP/MultiStateBmiCovid-19.

This study was approved by the Clinical Research Ethics Committee of the IDIAPJGol (project code: 20/070-PCV).

### Patient and public involvement

Participants of this study were not involved in setting the research question or the outcome measures, nor were they involved in the design or implementation of the study. No patients were asked to advise on interpretation or writing of results.

## Results

### Participants and observed outcomes

There were 4,765,757 adults from the SIDIAP population registered in the database on the 1st March 2020 (study index date) who were eligible to enter the study. We excluded 104,022 individuals due to having less than a year of prior clinical history; 306 due to having a prior COVID-19 clinical diagnosis or positive test; 41,588 due to being hospitalised or living in a nursing home on March 1st; 1,357,553 due to the unavailability of a BMI measurement; and 737,362 due to missing data on smoking status and/or the MEDEA deprivation index (Figure S1). A total of 2,524,926 participants were included in this study, of which 951,280 were living with underweight (2%) or normal weight (36%), 952,479 (38%) with overweight, and 621,167 (24%) with obesity (Table 1). The participants’ median BMI (interquartile [IQR] range) was 26 (24-30) kg/m^2^ and age was 52 (39-67) years. People living with underweight or normal weight were younger and more frequently female, current smokers, living in the least deprived areas of Catalonia and presenting with fewer comorbidities than people living with overweight or obesity (Table 1).

**Table 1.**
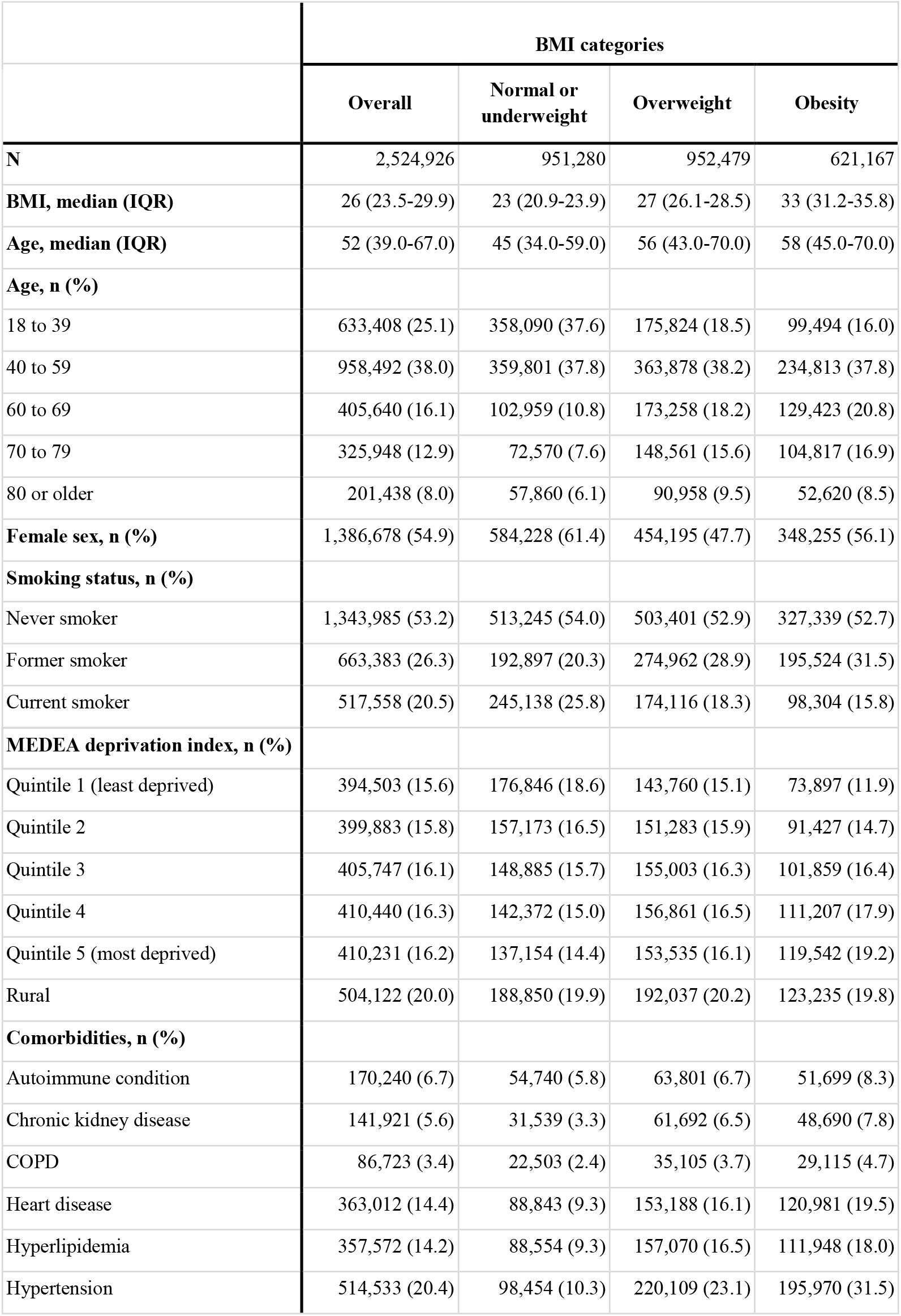
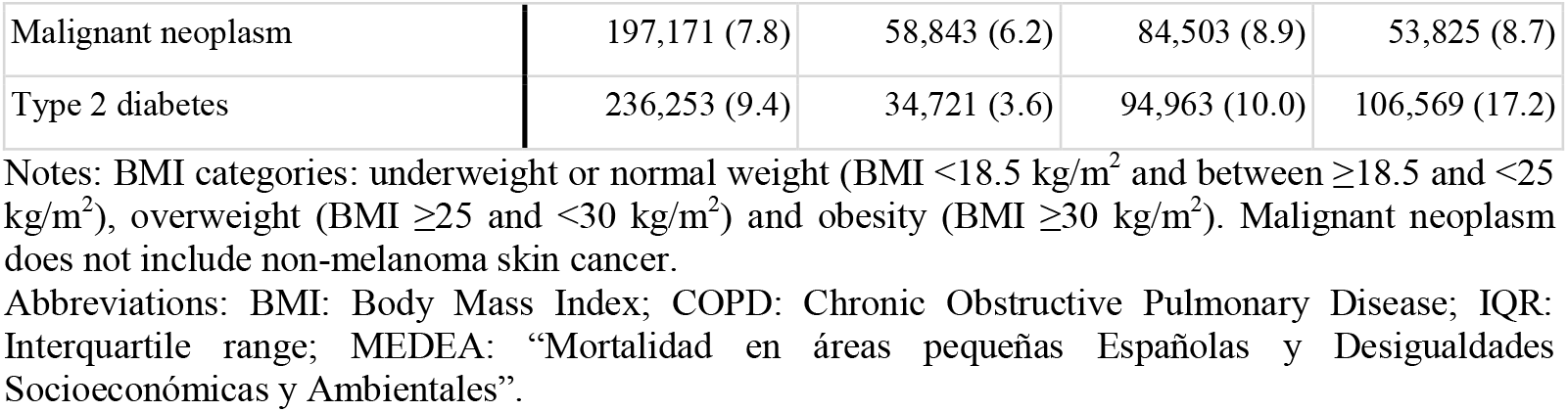
Descriptive statistics of the study population by body mass index categories.

|  | BMI categories |  |  |  |
| --- | --- | --- | --- | --- |
|  | Overall | Normal or<br>underweight | Overweight | Obesity |
| **N** | 2,524,926 | 951,280 | 952,479 | 621,167 |
| **BMI, median (IQR)** | 26 (23.5-29.9) | 23 (20.9-23.9) | 27 (26.1-28.5) | 33 (31.2-35.8) |
| **Age, median (IQR)** | 52 (39.0-67.0) | 45 (34.0-59.0) | 56 (43.0-70.0) | 58 (45.0-70.0) |
| **Age, n (%)** |  |  |  |  |
| 18 to 39 | 633,408 (25.1) | 358,090 (37.6) | 175,824 (18.5) | 99,494 (16.0) |
| 40 to 59 | 958,492 (38.0) | 359,801 (37.8) | 363,878 (38.2) | 234,813 (37.8) |
| 60 to 69 | 405,640 (16.1) | 102,959 (10.8) | 173,258 (18.2) | 129,423 (20.8) |
| 70 to 79 | 325,948 (12.9) | 72,570 (7.6) | 148,561 (15.6) | 104,817 (16.9) |
| 80 or older | 201,438 (8.0) | 57,860 (6.1) | 90,958 (9.5) | 52,620 (8.5) |
| **Female sex, n (%)** | 1,386,678 (54.9) | 584,228 (61.4) | 454,195 (47.7) | 348,255 (56.1) |
| **Smoking status, n (%)** |  |  |  |  |
| Never smoker | 1,343,985 (53.2) | 513,245 (54.0) | 503,401 (52.9) | 327,339 (52.7) |
| Former smoker | 663,383 (26.3) | 192,897 (20.3) | 274,962 (28.9) | 195,524 (31.5) |
| Current smoker | 517,558 (20.5) | 245,138 (25.8) | 174,116 (18.3) | 98,304 (15.8) |
| **MEDEA deprivation index, n (%)** |  |  |  |  |
| Quintile 1 (least deprived) | 394,503 (15.6) | 176,846 (18.6) | 143,760 (15.1) | 73,897 (11.9) |
| Quintile 2 | 399,883 (15.8) | 157,173 (16.5) | 151,283 (15.9) | 91,427 (14.7) |
| Quintile 3 | 405,747 (16.1) | 148,885 (15.7) | 155,003 (16.3) | 101,859 (16.4) |
| Quintile 4 | 410,440 (16.3) | 142,372 (15.0) | 156,861 (16.5) | 111,207 (17.9) |
| Quintile 5 (most deprived) | 410,231 (16.2) | 137,154 (14.4) | 153,535 (16.1) | 119,542 (19.2) |
| Rural | 504,122 (20.0) | 188,850 (19.9) | 192,037 (20.2) | 123,235 (19.8) |
| **Comorbidities, n (%)** |  |  |  |  |
| Autoimmune condition | 170,240 (6.7) | 54,740 (5.8) | 63,801 (6.7) | 51,699 (8.3) |
| Chronic kidney disease | 141,921 (5.6) | 31,539 (3.3) | 61,692 (6.5) | 48,690 (7.8) |
| COPD | 86,723 (3.4) | 22,503 (2.4) | 35,105 (3.7) | 29,115 (4.7) |
| Heart disease | 363,012 (14.4) | 88,843 (9.3) | 153,188 (16.1) | 120,981 (19.5) |
| Hyperlipidemia | 357,572 (14.2) | 88,554 (9.3) | 157,070 (16.5) | 111,948 (18.0) |
| Hypertension | 514,533 (20.4) | 98,454 (10.3) | 220,109 (23.1) | 195,970 (31.5) |
| Malignant neoplasm | 197,171 (7.8) | 58,843 (6.2) | 84,503 (8.9) | 53,825 (8.7) |
| Type 2 diabetes | 236,253 (9.4) | 34,721 (3.6) | 94,963 (10.0) | 106,569 (17.2) |

Notes: BMI categories: underweight or normal weight (BMI <18.5 kg/m<sup>2</sup> and between ≥18.5 and <25 kg/m<sup>2</sup>), overweight (BMI ≥25 and <30 kg/m<sup>2</sup>) and obesity (BMI ≥30 kg/m<sup>2</sup>). Malignant neoplasm does not include non-melanoma skin cancer.
Abbreviations: BMI: Body Mass Index; COPD: Chronic Obstructive Pulmonary Disease; IQR: Interquartile range; MEDEA: “Mortalidad en áreas pequeñas Españolas y Desigualdades Socioeconómicas y Ambientales”.

All the analysed baseline characteristics of the included individuals were meaningfully different (SMDs >0.1) from those of the excluded individuals due to missing information on BMI, smoking status and/or the MEDEA deprivation index (Table S1). Especially, the included participants were older (median age: 52 vs 44 years), more commonly female (55% vs 47%) and more frequently presenting with comorbidities (e.g., hypertension prevalence: 20% vs 8%).

After a median follow-up of 67 days of the initial (COVID-19 free) population, 57,443 (2.28%) were diagnosed with COVID-19 (median [IQR] BMI: 27 [24-30] kg/m^2^) and 5,191 (0.21%) were hospitalised without a prior outpatient diagnosis (29 [26-32] kg/m^2^) (Tables 2 and S2). Among the people diagnosed with COVID-19 in outpatient settings, 5,671 (10.62%) went on to be hospitalised (28 [26-32] kg/m^2^) and 1,166 (2.43%) died (27 [24-30] kg/m^2^) (median follow-up: 35 days). Finally, of the people that were hospitalised with COVID-19, 1,301 (19.22%) died (29 [26-32] kg/m^2^) (median follow-up: 37 days). The time at risk and absolute event rates of the participants by WHO categories of BMI are shown in Table 2 and the descriptive characteristics of people transitioning to each state of interest are available in Table S2.

**Table 2.** Time at risk, absolute event rates, and cumulative incidence over time by body mass index categories.

|  | From general population |  |  |  |  | From diagnosed with COVID-19 |  |  |  | From hospitalised with COVID-19 |  |  |
| --- | --- | --- | --- | --- | --- | --- | --- | --- | --- | --- | --- | --- |
|  |  |  | To diagnosis with COVID-19 | To hospitalised with COVID-19 | To death |  |  | To hospitalised with COVID-19 | To death |  |  | To death |
| BMI Categories | n | Follow-up in days, Median (min, IQR, max) | Events (cumulative incidence at 67 days) | Events (cumulative incidence at 67 days) | Events (cumulative incidence at 67 days) | n | Follow-up in days, Median (min, IQR, max) | Events (cumulative incidence at 45 days) | Events (cumulative incidence at 45 days) | n | Follow-up in days, Median (min, IQR, max) | Events (cumulative incidence at 45 days) |
| **Overall** | 2,524,926 | 67 (1, 67 to 67, 67) | 57,443 (2.28%) | 5,191 (0.21%) | 5,276 (0.21%) | 57,443 | 35 (0, 19 to 44, 66) | 5,671 (10.26%) | 1,166 (2.43%) | 10,862 | 37 (0, 27 to 43, 65) | 1,301 (19.22%) |
| **Normal or underweight** | 951,280 | 67 (1, 67 to 67, 67) | 20,931 (2.20%) | 939 (0.10%) | 1,907 (0.20%) | 20,931 | 36 (0, 21 to 44, 66) | 1,093 (5.47%) | 378 (2.15%) | 2,032 | 35 (0, 24 to 42, 65) | 269 (22.74%) |
| **Overweight** | 952,479 | 67 (1, 67 to 67, 67) | 21,369 (2.24%) | 2,233 (0.23%) | 2,024 (0.21%) | 21,369 | 35 (0, 17 to 44, 66) | 2,421 (11.76%) | 470 (2.64%) | 4,654 | 37 (0, 27 to 43, 65) | 541 (20.72%) |
| **Obesity** | 621,167 | 67 (1, 67 to 67, 67) | 15,143 (2.44%) | 2,019 (0.33%) | 1,345 (0.22%) | 15,143 | 34 (0, 15 to 43, 66) | 2,157 (14.76%) | 318 (2.51%) | 4,176 | 37 (0, 28 to 43, 65) | 491 (19.10%) |

Notes: BMI categories: underweight or normal weight (BMI <18.5 kg/m<sup>2</sup> and between ≥18.5 and <25 kg/m<sup>2</sup>), overweight (BMI ≥25 and <30 kg/m<sup>2</sup>) and obesity (BMI ≥30 kg/m<sup>2</sup>).
Abbreviations: BMI: Body Mass Index; COVID-19: Coronavirus Disease 2019; IQR: Interquartile range.

### Association between BMI and COVID-19 outcomes

The estimated shape of the association between BMI and COVID-19 outcomes for each transition, allowing for non-linearity, is shown in Figure 2. The fully adjusted analyses showed non-linear associations between BMI and risk of COVID-19 diagnosis, hospitalisation with COVID-19, and COVID-19 related death for all studied transitions (all p for non-linearity ≤0.001). The associations varied in shape and size by outcome of interest. Results for the crude and adjusted for age and sex models are shown in Figure S3 and Table S3. Interestingly, the shape of the fully adjusted models and those only adjusted for age and sex were similar.

**Figure 2.**
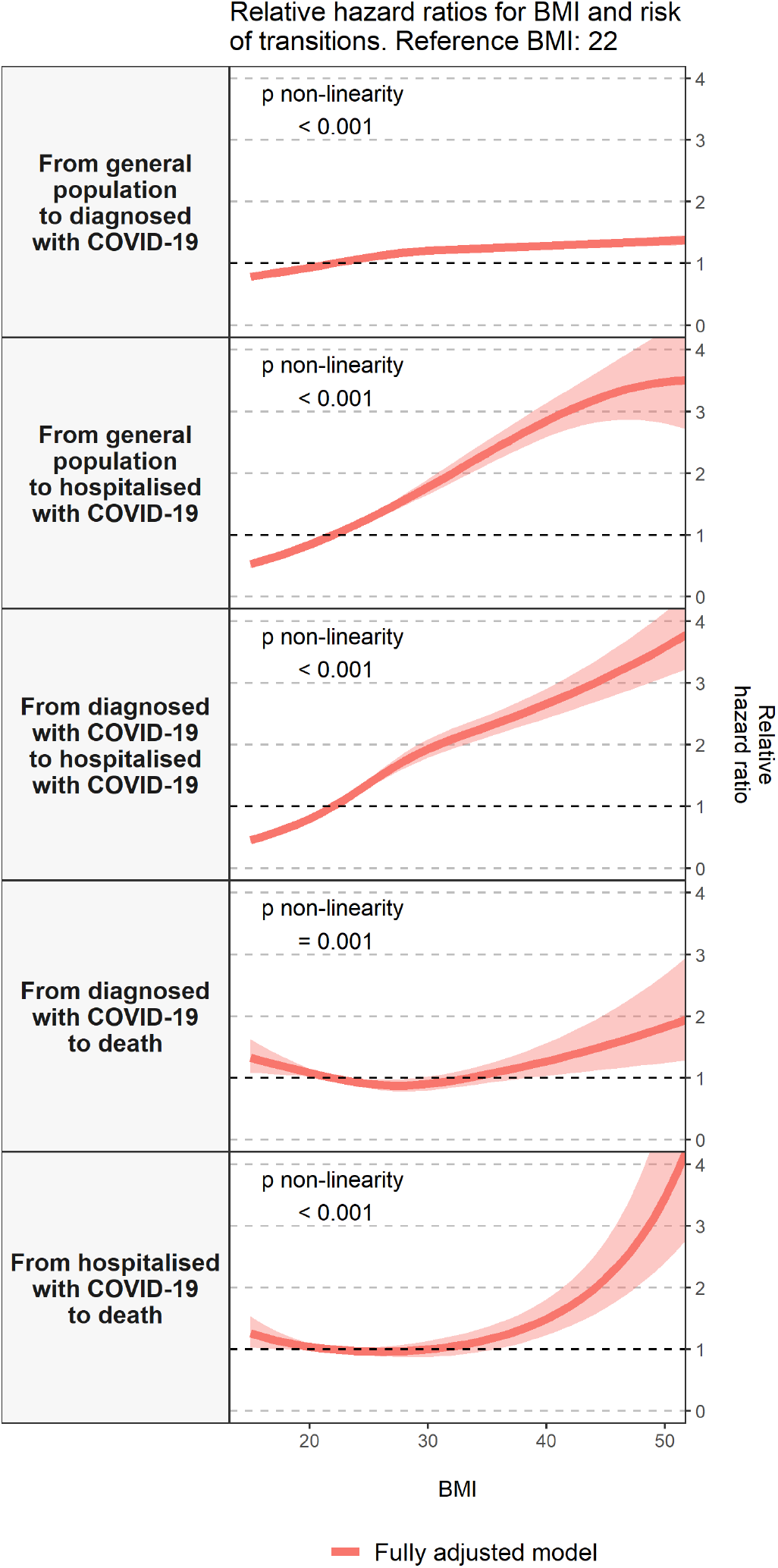
Association between body mass index and the risk of COVID-19 outcomes, allowing for non-linear effects, with 95% CIs. Notes: Models are adjusted for age, sex, smoking status and the MEDEA deprivation index Abbreviations: BMI: Body mass index; CI: Confidence interval; COVID-19: Coronavirus Disease 2019.

There was a modest positive association between BMI and the risk of COVID-19 *diagnosis* among the general population (Figure 2, Transition 1). Relative to a BMI of 22 kg/m^2^, the estimated hazard ratios were 0.81 (0.79-0.84) for someone with a BMI of 16 kg/m^2^; 1.10 (1.09-1.11) for a BMI of 25 kg/m^2^; 1.22 (1.19-1.24) for a BMI of 31 kg/m^2^, and 1.28 (1.25-1.32) for a BMI of 40 kg/m^2^ (HRs for these and other values of BMI are shown in Table 3).

**Table 3.** Hazards ratios of COVID-19 outcomes related to body mass index, with 95% CIs.

| BMI values (kg/m<sup>2</sup>) | From General Population |  | From diagnosed with COVID-19 |  | From hospitalised with COVID-19 |
| --- | --- | --- | --- | --- | --- |
|  | To diagnosed with COVID-19 | To hospitalised with COVID-19 | To hospitalised with COVID-19 | To death | To death |
| **16** | 0.81 (0.79-0.84) | 0.58 (0.53-0.64) | 0.51 (0.46-0.57) | 1.28 (1.07-1.52) | 1.20 (1.02-1.42) |
| **19** | 0.90 (0.89-0.91) | 0.77 (0.74-0.81) | 0.71 (0.68-0.75) | 1.13 (1.04-1.23) | 1.08 (1.00-1.16) |
| **22** | reference | reference | reference | reference | reference |
| **25** | 1.10 (1.09-1.11) | 1.27 (1.22-1.31) | 1.37 (1.31-1.43) | 0.90 (0.84-0.97) | 0.97 (0.91-1.03) |
| **28** | 1.17 (1.15-1.19) | 1.56 (1.47-1.66) | 1.74 (1.61-1.87) | 0.88 (0.78-0.99) | 0.97 (0.88-1.08) |
| **31** | 1.22 (1.19-1.24) | 1.88 (1.75-2.03) | 2.01 (1.86-2.18) | 0.93 (0.82-1.05) | 1.02 (0.89-1.17) |
| **34** | 1.24 (1.22-1.26) | 2.22 (2.04-2.41) | 2.22 (2.06-2.40) | 1.02 (0.89-1.17) | 1.11 (0.95-1.31) |
| **37** | 1.26 (1.23-1.29) | 2.54 (2.33-2.78) | 2.43 (2.24-2.64) | 1.14 (0.97-1.34) | 1.26 (1.06-1.51) |
| **40** | 1.28 (1.25-1.32) | 2.85 (2.58-3.13) | 2.66 (2.43-2.91) | 1.27 (1.03-1.56) | 1.49 (1.23-1.81) |
| **43** | 1.31 (1.26-1.36) | 3.11 (2.77-3.49) | 2.91 (2.62-3.23) | 1.42 (1.10-1.83) | 1.83 (1.47-2.29) |
| **47** | 1.34 (1.28-1.40) | 3.37 (2.87-3.96) | 3.27 (2.88-3.72) | 1.64 (1.18-2.27) | 2.56 (1.92-3.41) |
| **50** | 1.36 (1.29-1.44) | 3.48 (2.81-4.31) | 3.58 (3.09-4.15) | 1.82 (1.24-2.68) | 3.45 (2.41-4.96) |

Notes: Models are adjusted for age, sex, smoking status and the MEDEA deprivation index
Abbreviations: BMI: Body mass index; CI: Confidence interval; COVID-19: Coronavirus Disease 2019.

BMI was strongly associated with an increased risk of *hospitalisation* with COVID-19, either with or without a prior outpatient diagnosis (Figure 2, Transitions 2 and 3). Hazard ratios for hospitalisation without and with a prior diagnosis respectively, relative to a BMI of 22 kg/m^2^, were 0.58 (0.53-0.64) and 0.51 (0.46-0.57) for a BMI of 16 kg/m^2^; 1.27 (1.22-1.31) and 1.37 (1.31-1.43) for one of 25 kg/m^2^; 1.88 (1.75-2.03) and 2.01 (1.86-2.18) for one of 31 kg/m^2^; 2.85 (2.58-3.13) and 2.66 (2.43-2.91) for one of 40 kg/m^2^ (Table 3).

Finally, our analyses revealed an attenuated J-shaped association between BMI and risk of *death* either after an outpatient diagnosis or a hospitalisation with COVID-19 (Figure 2, Transitions 4 and 5). There was a modestly higher risk of death among individuals with very low BMIs but a higher risk for those with high BMIs. Relative to a BMI of 22 kg/m^2^, a BMI of 16 kg/m^2^ was associated with HRs of 1.28 (1.07-1.52) and 1.20 (1.02-1.42) for death after an outpatient diagnosis or a hospitalisation with COVID-19, respectively (Table 3). High BMIs became positively associated with death only at BMIs equal or greater than 37 kg/m^2^ among those that were previously hospitalised with COVID-19 (HR [95% CI]: 1.26 [1.06-1.51]) and 40 kg/m^2^ among those that were diagnosed with COVID-19 in outpatient settings (1.27 [1.03-1.56]).

### Effect modification by age and sex

There was evidence of effect modification by age and sex for two out of five studied transitions (p for interaction <0.0001) (Figure 3). Overall, the risk of COVID-19 outcomes related to increased BMI was higher for those aged between 18 and 59 years, compared to those in older age groups (Figure 3, Table S4). In addition, the effect of BMI on the risk of COVID-19 diagnosis and death after an outpatient diagnosis of COVID-19 differed for people aged 80 years or older compared to the other age groups. The risk of COVID-19 diagnosis for BMIs above 40 kg/m^2^ was higher for the oldest age group (HR, [95% CI]: 1.52 [1.33-1.74], relative to a BMI of 22 kg/m^2^) compared to those aged between 60 and 79 (1.10 [1.03-1.18]) or 59 years or younger (1.32 [1.28-1.37]) (Figure 3, Transition 1; Table S4 A). While there was no association between higher BMI and COVID-19 mortality after an outpatient diagnosis of COVID-19 for those in the oldest age group, there was a pronounced U-shaped association for those aged 59 years or younger and a J-shaped association for those aged between 60 and 79 years (Figure 3, Transition 4; Table S4 A). Associations were similarly shaped for females and males, although there was a significant interaction (p for interaction <0.0001) between BMI and sex for the first transition (males were at a slightly higher risk of being diagnosed with COVID-19 compared to females) (Figure 3, Transition 1; Table S4 B). Interestingly, the risk of death after hospitalisation with COVID-19 was stronger for females with BMIs above 43 kg/m^2^ (2.23 [1.66-3.00] relative to a BMI of 22 kg/m^2^) compared to males (1.30 [0.92-1.85]) (Figure 3, Transition 5; Table S4 B).

**Figure 3.**
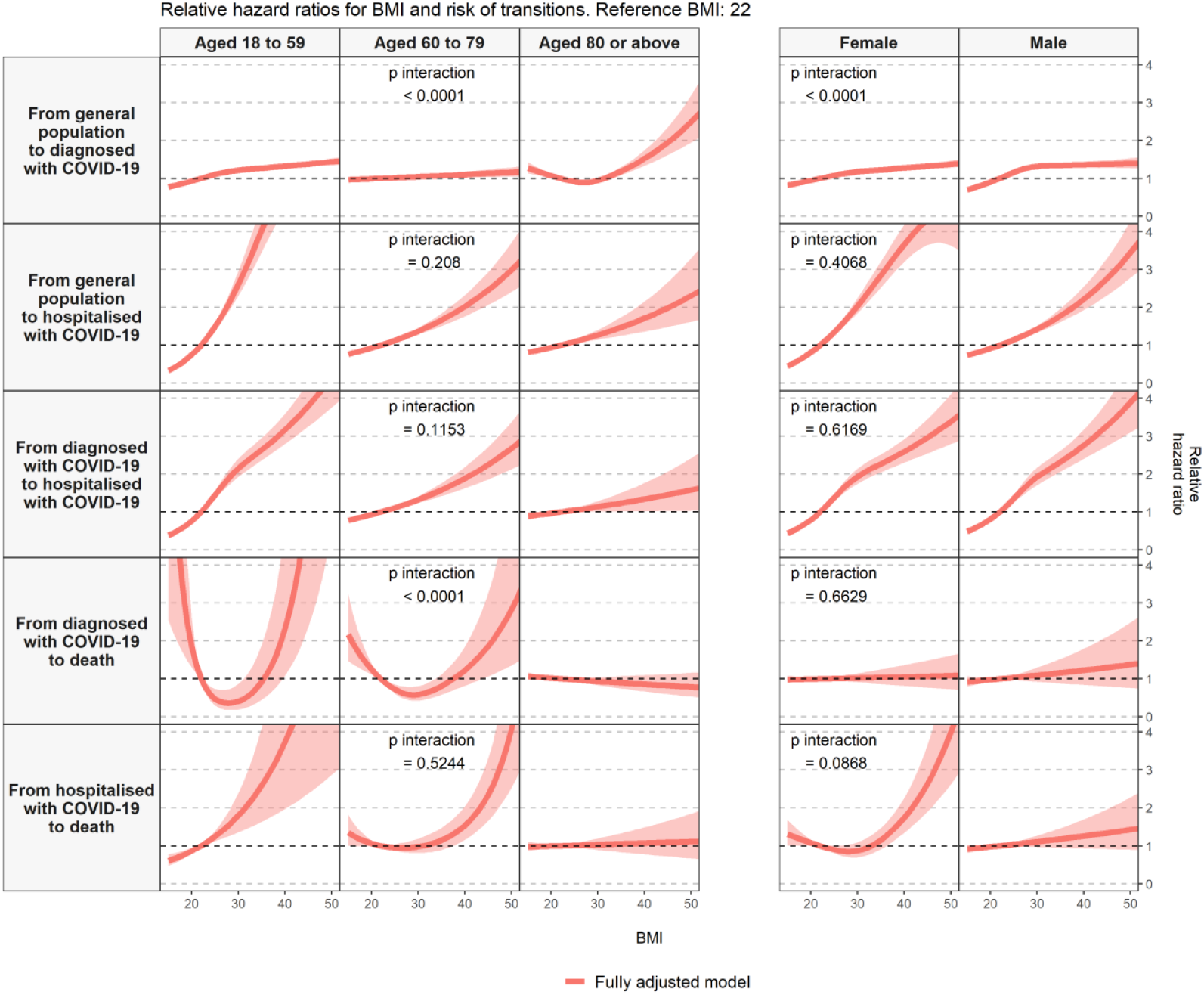
Effect modification by age and sex in the association between body mass index and the risk of COVID-19 outcomes, allowing for non-linear effects, with 95% CIs. Notes: Models are adjusted for age, sex, smoking status and the MEDEA deprivation index. Abbreviations: BMI: Body mass index; CI: Confidence interval; COVID-19: Coronavirus Disease 2019.

### Secondary and sensitivity analyses

The assumption of proportionality was violated for age in the first transition (*general populatio*n to a *diagnosed with COVID-19*). To account for this, we stratified the main model by calendar month although this appeared to have no meaningful impact on the studied associations. The risk of COVID-19 diagnosis related to increased BMI was slightly higher for those diagnosed in March compared to April (Figure S4, Table S5).

As a secondary analysis, we re-estimated the main models with BMI in WHO categories (Figure S5). Relative to the underweight and normal weight category of BMI, overweight and obesity were associated with a higher risk of being diagnosed with COVID-19 and hospitalised with COVID-19 (with and without a prior outpatient diagnosis of COVID-19). We did not observe an association between categorized BMI and risk of COVID-19 related death.

Our findings were robust to two sensitivity analyses. The shape of the studied associations and the estimated effect sizes of our main analyses were similar to those of the analyses in which did multiple imputations on missing data for BMI, smoking status, and the MEDEA deprivation index on one hand and in which we excluded BMI measurements older than five years on the other (Figures 2, S6-S7 and Tables 3, S6-S7).

## Discussion

### Principal findings

In this large cohort study that included 2,524,926 participants from the general population in Catalonia, Spain, we found that higher BMI was positively associated with risk of COVID-19 diagnosis, hospitalisation with COVID-19, and COVID-19 related death. The shape and effect size of the studied associations varied according to the COVID-19 outcome of interest as well as to individual factors such as age and sex. The relation between BMI and risk of COVID-19 related death was J-shaped with a modest positive risk of death among individuals with very low BMIs and a higher risk for those with higher BMI values. Overall, the associations between BMI and COVID-19 outcomes were stronger for those aged 59 years or younger compared to the older age groups, and similarly shaped among females and males, with specific exceptions.

### Strengths and weaknesses of this study

This study has several strengths. To our knowledge, this is the first longitudinal study to investigate the association between BMI and the course of the COVID-19 disease containing individual detailed BMI information and incident COVID-19 outcomes recorded in diverse healthcare settings from a large and representative population. The possibility to investigate COVID-19 trajectories in a single and sufficiently powered dataset, including systematic investigation of non-linearity and effect modification, is also a major strength. Further, the SIDIAP is representative of the Catalan general population, which suggests our findings are generalisable to Catalonia as well as to comparable regions. The results were robust when we explored the violation of the models’ assumptions, the possibility of selection bias due to missing data and exposure misclassification which is also a strong asset.

This study also has weaknesses. Firstly, this observational study only includes COVID-19 diagnoses of individuals who interacted with the health system. Especially in the first wave of the pandemic, testing was mainly restricted to severe cases of COVID-19. Although we aimed to reduce this bias by also including clinical diagnoses of COVID-19 (i.e., symptomatic individuals who were diagnosed with COVID-19 by general practitioners but were not confirmed by a positive test for SARS-CoV-2), we could not avoid missing asymptomatic or paucisymptomatic patients nor individuals who did not seek medical care. However, Catalonia has a tax-funded almost universal healthcare system. Secondly, we did not have the cause of death (only death after being diagnosed/hospitalised with COVID-19) which prevented us from attributing deaths to the disease. In the same direction, we will have missed individuals who died with COVID-19 but who were not identified as having been diagnosed or hospitalised with the disease. The likelihood of this outcome misclassification was probably reduced since we excluded individuals who were living in nursing homes at the beginning of the study. Thirdly, we did not have data on hospital visits that did not lead to an overnight stay nor admission to intensive services units during hospitalisation; this data can be useful to further study the progression of COVID-19 in detail. Fourthly, we did not have information on individual socioeconomic status nor the type of occupation of the participants; but we tried to minimize this limitation by including information on the MEDEA deprivation index. Fifthly, since individuals with high BMIs are more likely to have comorbidities, there could be a mediation of the association between BMI and COVID-19 outcomes by certain comorbidities, but exploring this topic was out of the scope of this study. Finally, the use of routinely collected data for research can raise concerns about data quality; however, BMI and COVID-19 data from the SIDIAP have previously been successfully repurposed for research.[27–29]

### Possible explanations

The increased risk of COVID-19 diagnosis among people with higher BMIs could be related to an increased vulnerability to SARS-CoV-2 and/or to higher exposure to the virus. Since obesity disproportionately affects disadvantaged populations, differential occupational risks (e.g., a higher likelihood of having manual occupations) should be explored in further studies.[2] The mechanisms by which higher BMI can increase COVID-19 severity include physical mechanisms (e.g., altered ventilation due to reduced diaphragm excursion), chronic inflammation and an impaired immune function.[6] A higher BMI is also a risk factor for several medical conditions that have been suggested to increase the risk of COVID-19 severity such as type 2 diabetes, COPD or heart disease (which were also more common in this study among patients with obesity compared to those with normal or underweight).[6,18] While part of the effect estimates of the present study might be mediated by some of these diseases, to disentangle these interrelations merits specific investigation. Other proposed explanations include delayed seek for medical care among individuals with obesity due to fear of being stigmatized (e.g., in this study, 26% of those diagnosed with COVID-19 and 39% of those hospitalised without an outpatient diagnosis of COVID-19 were living with obesity) as well as the difficulty of care in hospital settings for supportive therapies.[30,31]

### Research in context

Obesity, defined as a BMI ≥30kg/m^2^, has been consistently associated with the risk of testing positive for SARS-CoV-2 or being diagnosed with COVID-19.[6] Our dose-response analysis of BMI revealed that the risk of COVID-19 diagnosis increased linearly with higher values of BMI. Our findings are in line with a Mendelian randomization analysis which reported that genetically increased BMI was causally associated with testing positive for COVID-19.[32] In addition, a study among UK Biobank participants reported the association between BMI and risk of testing positive was similarly shaped to the one in our study.[8] These results highlight the importance of avoiding extremely high BMI cut-offs to determine vulnerable groups to the COVID-19 disease (e.g., the NHS only considers BMIs above 40 kg/m^2^ as risk groups for COVID-19).[17]

Our findings also revealed a much stronger association between BMI and COVID-19 diagnosis among those aged 80 years or older as compared to younger age groups and a modestly higher risk for males compared to females. While our findings are congruent with another study of the UK Biobank that analysed in-hospital SARS-CoV-2 test positivity regarding sex differences in risk, no effect modification by age group (70 years or older vs. younger than 70 years) was reported there.[33] The underlying age distribution of that subsample of the UK Biobank could explain this discrepancy; unfortunately, this information was not available for consultation.

Our findings of a strong positive association between BMI and risk of COVID-19 hospitalisation are in line with a meta-analysis of 19 studies that reported that obesity increases the odds of COVID-19 patients being hospitalised.[6] Our results suggest individuals with BMIs above 25 kg/m^2^ should be considered as a risk group for disease severity in the context of clinical management as well as policymaking.

While we did not find a statistically significant effect modification by age for associations between BMI and hospitalisation due to COVID-19, the HRs of COVID-19 were systematically higher for those aged 59 years or younger. These findings are in line with two hospital-based studies from the US. One reported a negative correlation between BMI and age among COVID-19 patients admitted to intensive care units in six US hospitals and another a positive association only among patients younger than 60 years of age compared to older adults.[34,35]

Two meta-analyses of 7 and 35 observational studies each, reported that obesity (BMI ≥30kg/m^2^), is associated with a higher risk of COVID-19 related mortality.[6,36] However, non-linear associations cannot be ignored in the field of BMI-related research, especially in relation to mortality.[4,5] Other large observational studies from the US and the UK using multiple categories of BMI, only found an association between morbid obesity (BMIs above 35 kg/m^2^ or 40 kg/m^2^) and COVID-19 mortality.[37–40] Our results for high BMIs are consistent with the latter studies, as our analyses revealed BMI was associated in a J-shaped fashion with the risk of COVID-19 related death: only BMIs above 37kg/m^2^ and 40kg/m^2^ were linked with a higher risk of death after a COVID-19 hospitalisation and after a COVID-19 outpatient diagnosis, respectively. Our findings were also aligned with those of a study conducted in a New York hospital which reported a J-shaped association between BMI and the risk of intubation or death.[7] Furthermore, our results provide important insights on the higher risk of COVID-19 related death for low BMIs (≤19kg/m^2^); while other studies also found this trend in their effect estimates, these were not significant, likely due to their smaller sample sizes.[7,39,40]

We also found that mortality risk related to an increased BMI was higher among individuals aged 69 years or younger compared to older adults. Four previous studies are much in line with our findings, while the opposite was reported in a meta-analysis.[7,33,36,39,40] Finally, we observed the risk of death after a hospitalisation with COVID-19 associated with BMI was higher among females, compared to males. The results of a study conducted among UK Biobank participants were congruent with our findings, while one performed in a New York hospital found a higher risk among males and others found opposite or null differences by sex.[7,33,36,39,40] Further studies are recommended to investigate this aspect.

### Meaning of the study and future research

In this large cohort study, we provide a comprehensive analysis of the association between BMI and the course of the COVID-19 disease during the first wave of the pandemic in Catalonia, Spain. We explored non-linearity and effect modification by age and sex based on the results of previous studies investigating associations between BMI and multiple health outcomes. Our analyses revealed that BMI is positively associated with being diagnosed and hospitalised with COVID-19, and in a J-shaped fashion with the risk of COVID-19 related death. Furthermore, the risk of COVID-19 outcomes related to increased BMI, is higher for individuals aged 59 years or younger, compared to older people. In light of these findings and the global health emergency context, individuals with a high BMI (especially those in younger age groups) should be targeted in preventive strategies and prioritized in clinical practice. More research is needed to unravel the mechanisms underlying these relationships as well as to determine to what extent these associations are mediated by intermediate factors such as specific comorbidities. Furthermore, second wave-updated data and longer follow-ups are necessary to provide a better understanding of the effect of BMI on the progression of the COVID-19 disease.

## Data Availability

In accordance with current European and national law, the data used in this study is only available for the researchers participating in this study. Thus, we are not allowed to distribute or make publicly available the data to other parties. However, researchers from public institutions can request data from SIDIAP if they comply with certain requirements. Further information is available online (https://www.sidiap.org/index.php/menu-solicitudesen/application-proccedure) or by contacting Anna Moleras.

## Acknowledgements

We would like to acknowledge the patients who suffered from or died of this devastating disease, and their families and carers. We would also like to thank the healthcare professionals involved in the management of COVID-19 during these challenging times, from primary care to intensive care units in the Catalan healthcare system.

## Transparency statement

MR and TDS as guarantors of the manuscript affirm that the manuscript is an honest, accurate, and transparent account of the study being reported; that no important aspects of the study have been omitted; and that any discrepancies from the study as planned have been explained.

## Competing interests

All authors have completed the ICMJE uniform disclosure form at www.icmje.org/coi_disclosure.pdf and declare: no support from any organisation for the submitted work; Dr Prieto-Alhambra reports grants and other from AMGEN; grants, non-financial support and other from UCB Biopharma; grants from Les Laboratoires Servier, outside the submitted work; and Janssen, on behalf of IMI-funded EHDEN and EMIF consortiums, and Synapse Management Partners have supported training programs organized by DPA’s department and open for external participants. No other relationships or activities that could appear to have influenced the submitted work.

## Funding and role of the funding source

This project is funded by the Health Department from the Generalitat de Catalunya with a grant for research projects on SARS-CoV-2 and COVID-19 disease organized by the Direcció General de Recerca i Innovació en Salut. This project has received support from the European Health Data and Evidence Network (EHDEN) project. EHDEN received funding from the Innovative Medicines Initiative 2 Joint Undertaking (JU) under grant agreement No 806968. The JU receives support from the European Union’s Horizon 2020 research and innovation programme and EFPIA. The University of Oxford received a grant related to this work from the Bill & Melinda Gates Foundation (Investment ID INV-016201), and partial support from the UK National Institute for Health Research (NIHR) Oxford Biomedical Research Centre. DPA is funded through a National Institute for Health Research (NIHR) Senior Research Fellowship (Grant number SRF-2018-11-ST2-004). The views expressed in this publication are those of the authors and not necessarily those of the NHS, the National Institute for Health Research or the Department of Health. The funders had no role in study design, data collection, and analysis, decision to publish, or preparation of the manuscript.

## Author contributions

SFB, MA, TDS mapped source data to the OMOP-CDM. MR, AP, EB and TDS led the data analysis. MR and HF performed a literature review. All authors were involved in the study conception and design, interpretation of the results, and the preparation of the manuscript.

## Disclaimer

Where authors are identified as personnel of the International Agency for Research on Cancer / World Health Organization, the authors alone are responsible for the views expressed in this article and they do not necessarily represent the decisions, policy or views of the International Agency for Research on Cancer / World Health Organization.

## Abbreviations

BIC: Bayesian Information Criterion
BMI: body mass index
CDM: Common Data Model
CI: confidence interval
COPD: chronic obstructive pulmonary disease
COVID-19: coronavirus disease 2019
EHR: Electronic Health Record
HR: hazard ratio
ICD-10-CM: International Classification of Diseases, Tenth Revision, Clinical Modification
IQR: Interquartile Range
MEDEA: Mortalidad en áreas pequeñas Españolas y Desigualdades Socioeconómicas y Ambientales
OMOP: Observational Medical Outcomes Partnership
OHDSI: Observational Health Data Sciences and Informatics
RT-PCR: Reverse Transcription Polymerase Chain Reaction
SARS-CoV-2: Severe Acute Respiratory Syndrome Coronavirus 2
SIDIAP: Information System for Research in Primary Care
SMD: Standardized Mean Difference
WHO: World Health Organization.

