## Supplementary Material for "Body mass index and risk of COVID-19 diagnosis, hospitalisation, and death: a population-based multi-state cohort analysis including 2,524,926 people in Catalonia, Spain"

Martina Recalde, *PhD student*[1,2], Andrea Pistillo, *statistician*[1], Sergio Fernandez-Bertolin, *data scientist*[1], Elena Roel, *PhD student*[1,2], Maria Aragon, *SIDIAP coordinator*[1], Heinz Freisling, *scientist*[3], Daniel Prieto-Alhambra, *Professor of pharmaco- and device epidemiology*[4], Edward Burn, *post-doctoral researcher*[1,4]*, Talita Duarte-Salles, *senior epidemiologist*[1]*

*Joint senior authorship

1. Fundació Institut Universitari per a la recerca a l'Atenció Primària de Salut Jordi Gol i Gurina (IDIAPJGol), Barcelona, Spain
2. Universitat Autònoma de Barcelona, Spain
3. International Agency for Research on Cancer (IARC-WHO), 150 Cours Albert Thomas, 69008 Lyon, France
4. Centre for Statistics in Medicine, NDORMS, University of Oxford

**CORRESPONDING AUTHORS**
Daniel Prieto-Alhambra
Botnar Research Centre, Windmill Road
OX37LD, Oxford, United Kingdom


Talita Duarte-Salles
Fundació Institut Universitari per a la recerca a l'Atenció Primària de Salut Jordi Gol i Gurina (IDIAPJGol)
Gran Via Corts Catalanes, 587 àtic
08007 Barcelona - Spain


### Appendix

**Figure S1.** Flowchart with the inclusion and exclusion criteria of the study population

**Figure S2.** Directed Acyclic Graph for the possible causal effect of obesity on COVID-19 outcomes used to adjust the Cox proportional hazard models

**Table S1.** Characteristics of individuals excluded from the study due to missing information on BMI, smoking status and/or the MEDEA deprivation index

**Table S2.** Characteristics of the study participants by multi-state transition

**Figure S3.** Association between body mass index and the risk of COVID-19 outcomes with different types of model-adjustment, allowing for non-linear effects, with 95% CIs

**Table S3.** Hazards ratios of COVID-19 outcomes related to body mass index, with 95% CIs, for the unadjusted, adjusted for age and sex and fully adjusted models

**Table S4.** Hazards ratios of COVID-19 outcomes related to body mass index, with 95% CIs, stratified by age and sex

**Figure S4**. Association between body mass index and the risk of COVID-19 diagnosis, allowing for non-linear effects, with 95% CIs, stratified by calendar month

**Table S5.** Hazard Ratios of the risk of COVID-19 diagnosis related to body mass index, allowing for non-linear effects, with 95% CIs, stratified by calendar month

**Figure S5.** Hazard Ratios of COVID-19 outcomes related to body mass index in categories, with 95% CIs

**Figure S6.** Association between body mass index and the risk of COVID-19 outcomes, allowing for non-linear effects, with 95% CIs, after multiple imputation of missing data on BMI, smoking status, and/or the MEDEA deprivation index

**Table S6.** Hazards ratios of COVID-19 outcomes related to body mass index, with 95% CIs, after multiple imputation of missing data on BMI, smoking status, and/or the MEDEA deprivation index

**Figure S7.** Association between body mass index and the risk of COVID-19 outcomes, allowing for non-linear effects, with 95% CIs among people with a BMI measurement not older than five years prior to the index date

**Table S7.** Hazards ratios of COVID-19 outcomes related to body mass index, with 95% CIs among people with a BMI measurement not older than five years prior to the index date

**Figure S1.** Flowchart with the inclusion and exclusion criteria of the study population

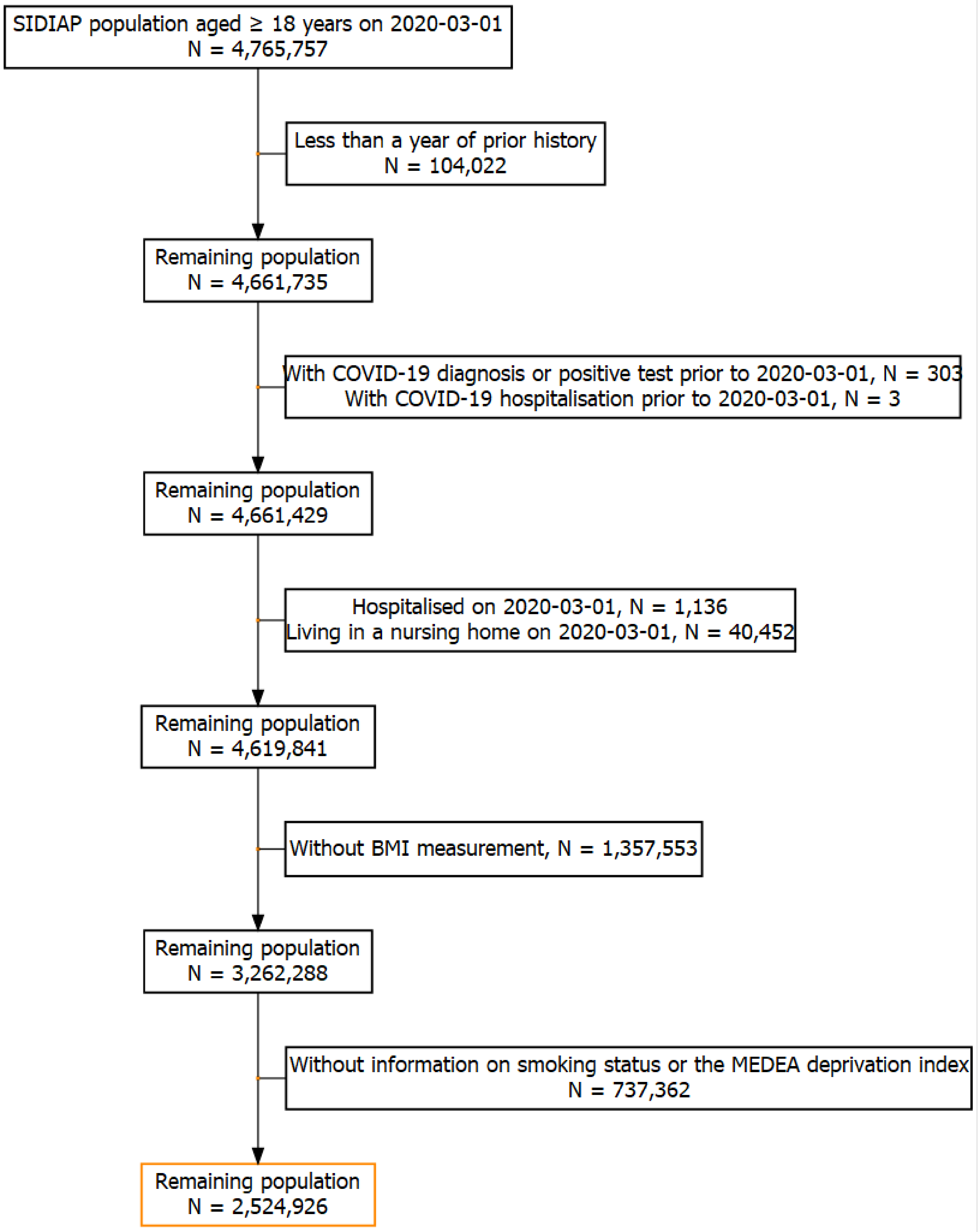


Abbreviations: BMI: Body Mass Index; COVID-19: Coronavirus Disease 2019; MEDEA: "Mortalidad en áreas pequeñas Españolas y Desigualdades Socioeconómicas y Ambientales"; SIDIAP: Information System for Research in Primary Care.

**Figure S2.** Directed Acyclic Graph for the possible causal effect of obesity on COVID-19 outcomes used to adjust the Cox proportional hazard models

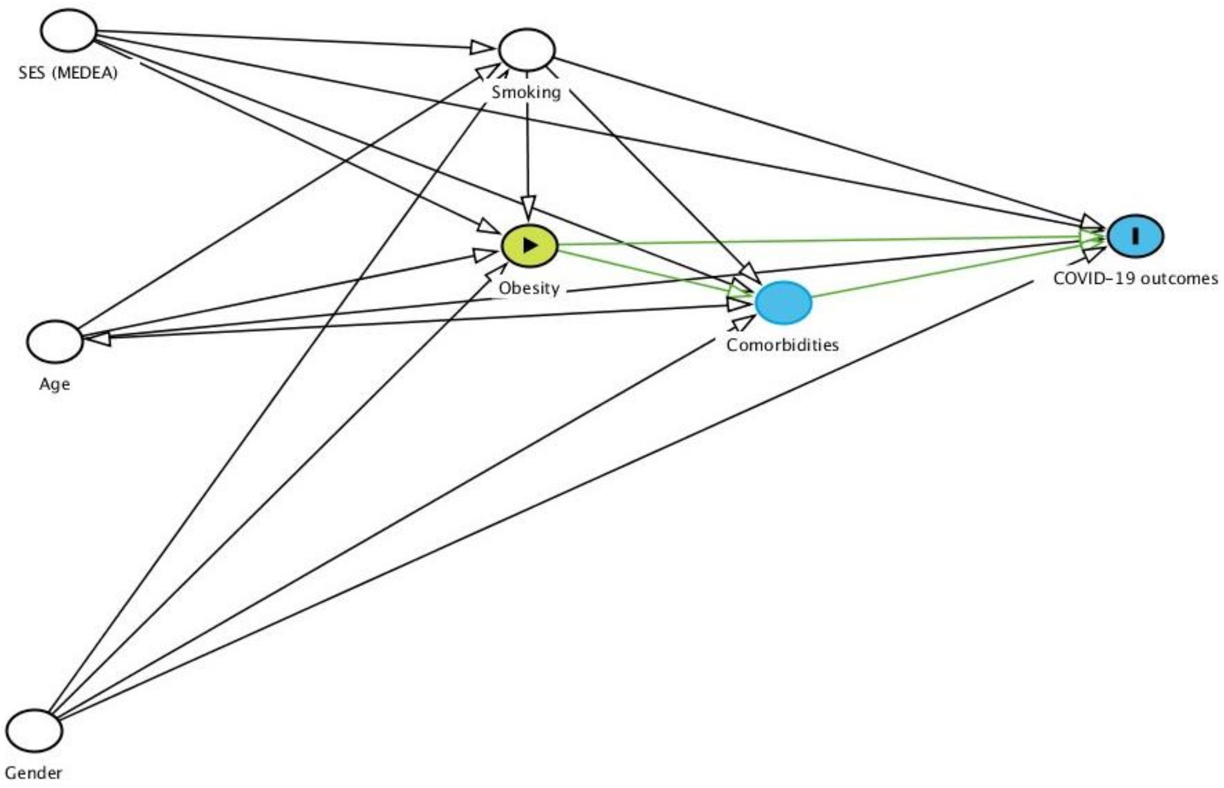


Notes: Green lines indicate a possible pathway between the exposure and outcome of interest.
Abbreviations: COVID-19: Coronavirus Disease 2019; MEDEA: "Mortalidad en áreas pequeñas Españolas y Desigualdades Socioeconómicas y Ambientales"; SES: Socioeconomic status.

**Table S1.** Characteristics of individuals excluded from the study due to missing information on BMI, smoking status and/or the MEDEA deprivation index

| | Overall | BMI recorded and complete data on covariates | Missing data on BMI[a] or covariates[b] | SMD | Missing BMI[a] | BMI recorded but missing data on covariates[b] |
|---|---|---|---|---|---|---|
| **N** | 4,619,841 | 2,524,926 | 2,094,915 | | 1,357,553 | 737,362 |
| **BMI, median (IQR)** | 26.5 (23.5-30.0) | 26.5 (23.5-29.9) | 26.6 (23.6-30.1) | 0.260 | - | 26.6 (23.6-30.1) |
| **Age, median (IQR)** | 48 (36.0-63.0) | 52 (39.0-67.0) | 44 (32.0-57.0) | 0.394 | 40 (27.0-50.0) | 56 (43.0-71.0) |
| **Age, n (%)** | | | | 0.371 | | |
| 18 to 39 | 1,437,297 (31.1) | 633,408 (25.1) | 803,889 (38.4) | | 678,297 (50.0) | 125,592 (17.0) |
| 40 to 59 | 1,785,852 (38.7) | 958,492 (38.0) | 827,360 (39.5) | | 535,200 (39.4) | 292,160 (39.6) |
| 60 to 69 | 615,538 (13.3) | 405,640 (16.1) | 209,898 (10.0) | | 94,137 (6.9) | 115,761 (15.7) |
| 70 to 79 | 468,662 (10.1) | 325,948 (12.9) | 142,714 (6.8) | | 32,370 (2.4) | 110,344 (15.0) |
| 80 or older | 312,492 (6.8) | 201,438 (8.0) | 111,054 (5.3) | | 17,549 (1.3) | 93,505 (12.7) |
| **Female sex, n (%)** | 2,361,933 (51.1) | 1,386,678 (54.9) | 975,255 (46.6) | 0.168 | 545,827 (40.2) | 429,428 (58.2) |
| **Smoking status, n (%)** | | | | 1.841 | | |
| Never smoker | 712918 (15.4) | 517558 (20.5) | 195360 (9.3) | | 159987 (11.8) | - |
| Former smoker | 773325 (16.7) | 663383 (26.3) | 109942 (5.2) | | 66878 (4.9) | - |
| Current smoker | 1835150 (39.7) | 1343985 (53.2) | 491165 (23.4) | | 406087 (29.9) | - |
| Missing | 1298448 (28.1) | - | 1298448 (62.0) | | 724601 (53.4) | - |
| **MEDEA deprivation index, n (%)** | | | | 0.574 | | |
| Quintile 1 (least deprived) | 714440 (15.5) | 394503 (15.6) | 319937 (15.3) | | 248387 (18.3) | - |
| Quintile 2 | 704177 (15.2) | 399883 (15.8) | 304294 (14.5) | | 215825 (15.9) | - |

| | | | | | | |
|---|---|---|---|---|---|---|
| Quintile 3 | 697299 (15.1) | 405747 (16.1) | 291552 (13.9) | | 201562 (14.8) | - |
| Quintile 4 | 693046 (15.0) | 410440 (16.3) | 282606 (13.5) | | 189614 (14.0) | - |
| Quintile 5 (most deprived) | 687266 (14.9) | 410231 (16.2) | 277035 (13.2) | | 189495 (14.0) | - |
| Rural | 832483 (18.0) | 504122 (20.0) | 328361 (15.7) | | 219066 (16.1) | - |
| Missing | 291130 (6.3) | - | 291130 (13.9) | | 93604 (6.9) | - |
| **Comorbidities, n (%)** | | | | | | |
| Autoimmune condition | 259,383 (5.6) | 170,240 (6.7) | 89,143 (4.3) | 0.109 | 36,253 (2.7) | 52,890 (7.2) |
| Chronic kidney disease | 201,456 (4.4) | 141,921 (5.6) | 59,535 (2.8) | 0.138 | 6,006 (0.4) | 53,529 (7.3) |
| COPD | 119,669 (2.6) | 86,723 (3.4) | 32,946 (1.6) | 0.119 | 4,112 (0.3) | 28,834 (3.9) |
| Heart disease | 516,553 (11.2) | 363,012 (14.4) | 153,541 (7.3) | 0.228 | 31,384 (2.3) | 122,157 (16.6) |
| Hyperlipidemia | 505,365 (10.9) | 357,572 (14.2) | 147,793 (7.1) | 0.232 | 43,730 (3.2) | 104,063 (14.1) |
| Hypertension | 687,824 (14.9) | 514,533 (20.4) | 173,291 (8.3) | 0.351 | 35,457 (2.6) | 137,834 (18.7) |
| Malignant neoplasm | 289,827 (6.3) | 197,171 (7.8) | 92,656 (4.4) | 0.142 | 27,211 (2.0) | 65,445 (8.9) |
| Type 2 diabetes | 317,254 (6.9) | 236,253 (9.4) | 81,001 (3.9) | 0.222 | 6,025 (0.4) | 74,976 (10.2) |

Notes: a) Includes individuals who do not have a BMI recorded in their electronic health records and those with a BMI measurement recorded before being 18 years of age. b) Includes individuals without information on smoking or socioeconomic status. Malignant neoplasm does not include non-melanoma skin cancer.

Abbreviations: BMI: Body Mass Index; COPD: Chronic Obstructive Pulmonary Disease; IQR: Interquartile range; MEDEA: “Mortalidad en áreas pequeñas Españolas y Desigualdades Socioeconómicas y Ambientales”; SMD: Standardized mean difference.

**Table S2.** Characteristics of the study participants by multi-state transition

| | General population | From general population | | | From diagnosed with COVID-19 | | From hospitalised with COVID-19 |
|---|---|---|---|---|---|---|---|
| | | To diagnosed with COVID-19 | To hospitalised with COVID-19 | To death | To hospitalised with COVID-19 | To death | To death |
| N | 2,524,926 | 57,443 | 5,191 | 5,276 | 5,671 | 1,166 | 1,301 |
| **BMI, median (IQR)** | 26 (23.5-29.9) | 27 (23.5-30.3) | 29 (25.9-32.1) | 27 (23.6-30.1) | 28 (25.7-32.0) | 27 (24.1-30.4) | 29 (25.6-31.9) |
| **BMI in WHO categories, n (%)** | | | | | | | |
| Normal or underweight | 951,280 (37.7) | 20,931 (36.4) | 939 (18.1) | 1,907 (36.1) | 1,093 (19.3) | 378 (32.4) | 269 (20.7) |
| Overweight | 952,479 (37.7) | 21,369 (37.2) | 2,233 (43.0) | 2,024 (38.4) | 2,421 (42.7) | 470 (40.3) | 541 (41.6) |
| Obesity | 621,167 (24.6) | 15,143 (26.4) | 2,019 (38.9) | 1,345 (25.5) | 2,157 (38.0) | 318 (27.3) | 491 (37.7) |
| **Age, median (IQR)** | 52 (39.0-67.0) | 48 (38.0-60.0) | 70 (58.0-79.0) | 85 (75.0-90.0) | 60 (50.0-72.0) | 86 (79.0-91.0) | 79 (72.0-86.0) |
| **Age, n (%)** | | | | | | | |
| 18 to 39 | 633,408 (25.1) | 16,502 (28.7) | 264 (5.1) | 19 (0.4) | 505 (8.9) | 1 (0.1) | 5 (0.4) |
| 40 to 59 | 958,492 (38.0) | 26,455 (46.1) | 1,140 (22.0) | 295 (5.6) | 2,203 (38.8) | 29 (2.5) | 69 (5.3) |
| 60 to 69 | 405,640 (16.1) | 6,927 (12.1) | 1,100 (21.2) | 526 (10.0) | 1,176 (20.7) | 69 (5.9) | 163 (12.5) |
| 70 to 79 | 325,948 (12.9) | 4,198 (7.3) | 1,490 (28.7) | 974 (18.5) | 1,083 (19.1) | 195 (16.7) | 446 (34.3) |
| 80 or older | 201,438 (8.0) | 3,361 (5.9) | 1,197 (23.1) | 3,462 (65.6) | 704 (12.4) | 872 (74.8) | 618 (47.5) |
| **Female sex, n (%)** | 1,386,678 (54.9) | 35,236 (61.3) | 2,168 (41.8) | 2,379 (45.1) | 2,568 (45.3) | 632 (54.2) | 471 (36.2) |
| **Smoking status, n (%)** | | | | | | | |
| Never smoker | 517,558 (20.5) | 10,103 (17.6) | 328 (6.3) | 509 (9.6) | 346 (6.1) | 64 (5.5) | 58 (4.5) |

| | | | | | | | |
|---|---|---|---|---|---|---|---|
| Former smoker | 663,383 (26.3) | 15,886 (27.7) | 2,174 (41.9) | 2,268 (43.0) | 2,103 (37.1) | 502 (43.1) | 635 (48.8) |
| Current smoker | 1,343,985 (53.2) | 31,454 (54.8) | 2,689 (51.8) | 2,499 (47.4) | 3,222 (56.8) | 600 (51.5) | 608 (46.7) |
| **MEDEA deprivation index, n (%)** | | | | | | | |
| Quintile 1 (least deprived) | 394,503 (15.6) | 8,755 (15.2) | 759 (14.6) | 873 (16.5) | 783 (13.8) | 239 (20.5) | 200 (15.4) |
| Quintile 2 | 399,883 (15.8) | 9,423 (16.4) | 835 (16.1) | 750 (14.2) | 943 (16.6) | 162 (13.9) | 191 (14.7) |
| Quintile 3 | 405,747 (16.1) | 9,624 (16.8) | 909 (17.5) | 736 (13.9) | 1,019 (18.0) | 189 (16.2) | 228 (17.5) |
| Quintile 4 | 410,440 (16.3) | 10,077 (17.5) | 977 (18.8) | 690 (13.1) | 1,037 (18.3) | 111 (9.5) | 219 (16.8) |
| Quintile 5 (most deprived) | 410,231 (16.2) | 9,942 (17.3) | 1,018 (19.6) | 644 (12.2) | 1,027 (18.1) | 106 (9.1) | 190 (14.6) |
| Rural | 504,122 (20.0) | 9,622 (16.8) | 693 (13.4) | 1,583 (30.0) | 862 (15.2) | 359 (30.8) | 273 (21.0) |
| **Comorbidities, n (%)** | | | | | | | |
| Autoimmune condition | 170,240 (6.7) | 4,025 (7.0) | 541 (10.4) | 615 (11.7) | 496 (8.7) | 125 (10.7) | 152 (11.7) |
| Chronic kidney disease | 141,921 (5.6) | 2,500 (4.4) | 890 (17.1) | 1,783 (33.8) | 585 (10.3) | 402 (34.5) | 404 (31.1) |
| COPD | 86,723 (3.4) | 1,622 (2.8) | 513 (9.9) | 717 (13.6) | 326 (5.7) | 126 (10.8) | 179 (13.8) |
| Heart disease | 363,012 (14.4) | 7,083 (12.3) | 1,674 (32.2) | 2,733 (51.8) | 1,228 (21.7) | 561 (48.1) | 625 (48.0) |
| Hyperlipidemia | 357,572 (14.2) | 7,439 (13.0) | 1,040 (20.0) | 786 (14.9) | 1,036 (18.3) | 184 (15.8) | 260 (20.0) |
| Hypertension | 514,533 (20.4) | 9,923 (17.3) | 1,726 (33.2) | 1,940 (36.8) | 1,609 (28.4) | 422 (36.2) | 481 (37.0) |
| Malignant neoplasm | 197,171 (7.8) | 3,588 (6.2) | 861 (16.6) | 1,828 (34.6) | 641 (11.3) | 272 (23.3) | 326 (25.1) |
| Type 2 diabetes | 236,253 (9.4) | 4,327 (7.5) | 1,153 (22.2) | 1,313 (24.9) | 927 (16.3) | 288 (24.7) | 386 (29.7) |

Notes: BMI categories: underweight or normal weight (BMI <18.5 kg/m$^2$ and between ≥18.5 and <25 kg/m$^2$), overweight (BMI ≥25 and <30 kg/m$^2$) and obesity (BMI ≥30 kg/m$^2$). Malignant neoplasm does not include non-melanoma skin cancer.

Abbreviations: BMI: Body Mass Index; COPD: Chronic Obstructive Pulmonary Disease; COVID-19: Coronavirus Disease 2019; IQR: Interquartile range; MEDEA: "Mortalidad en áreas pequeñas Españolas y Desigualdades Socioeconómicas y Ambientales"; WHO: World Health Organization.

**Figure S3.** Association between body mass index and the risk of COVID-19 outcomes with different types of model-adjustment, allowing for non-linear effects, with 95% CIs

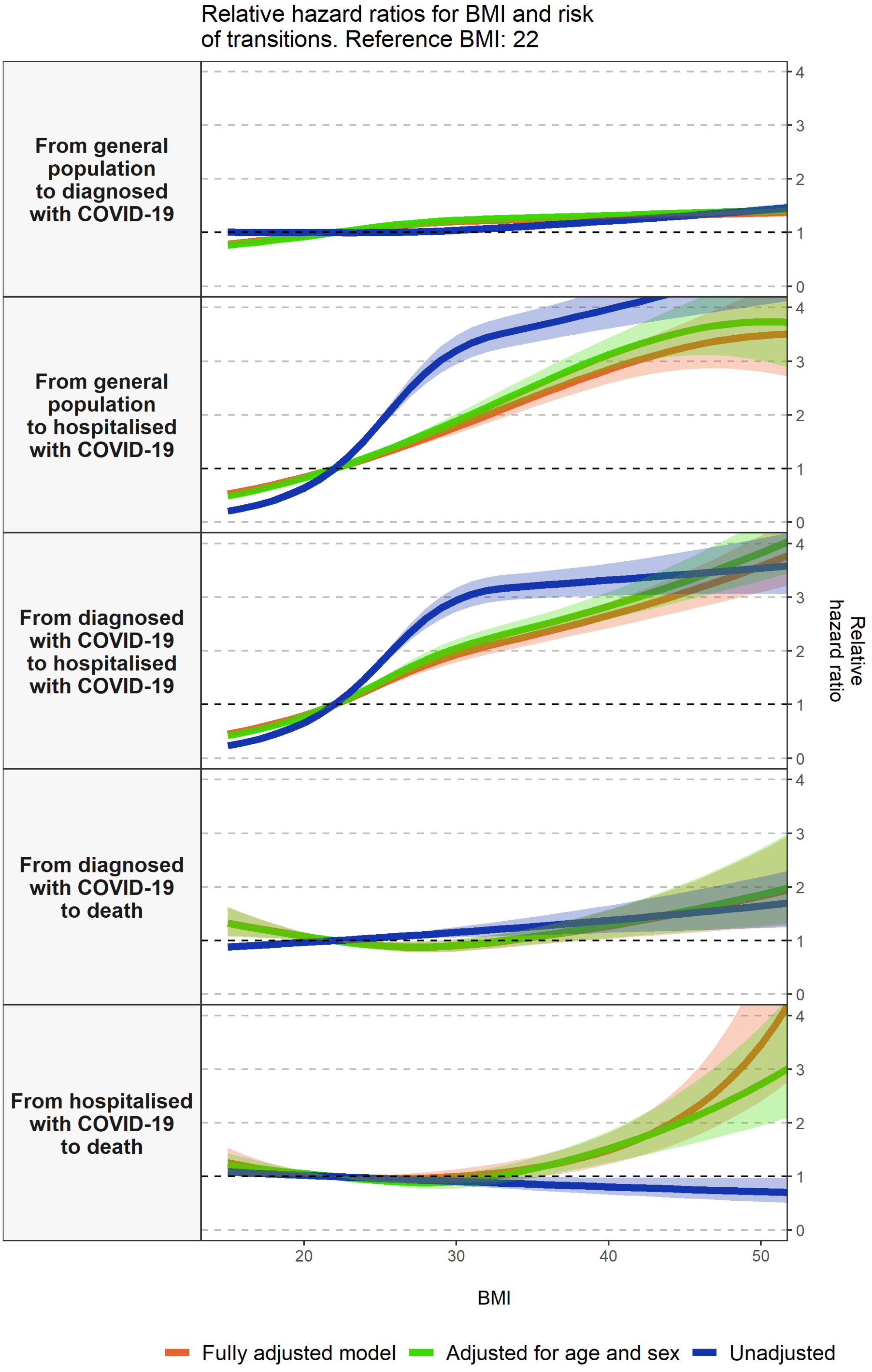


Notes: the fully adjusted models are adjusted for age, sex, smoking status and the MEDEA deprivation index.
Abbreviations: BMI: Body mass index; CI: Confidence interval; COVID-19: Coronavirus Disease 2019.

**Table S3.** Hazards ratios of COVID-19 outcomes related to body mass index, with 95% CIs, for the unadjusted, adjusted for age and sex and fully adjusted models

| | **From general population** | | | | | | **From diagnosed with COVID-19** | | | | | | **From hospitalised with COVID-19** | | |
|---|---|---|---|---|---|---|---|---|---|---|---|---|---|---|---|
| | **to diagnosed with COVID-19** | | | **to hospitalised with COVID-19** | | | **to hospitalised with COVID-19** | | | **to death** | | | **to death** | | |
| **BMI values ($kg/m^2$)** | **Un-adjusted** | **Adjusted for age and sex** | **Fully adjusted** | **Un-adjusted** | **Adjusted for age and sex** | **Fully adjusted** | **Un-adjusted** | **Adjusted for age and sex** | **Fully adjusted** | **Un-adjusted** | **Adjusted for age and sex** | **Fully adjusted** | **Un-adjusted** | **Adjusted for age and sex** | **Fully adjusted** |
| **16** | 1.01 (0.98-1.03) | 0.80 (0.78-0.82) | 0.81 (0.79-0.84) | 0.26 (0.23-0.29) | 0.54 (0.49-0.60) | 0.58 (0.53-0.64) | 0.29 (0.26-0.32) | 0.48 (0.43-0.53) | 0.51 (0.46-0.57) | 0.90 (0.85-0.96) | 1.27 (1.07-1.51) | 1.28 (1.07-1.52) | 1.07 (1.01-1.14) | 1.17 (1.01-1.36) | 1.20 (1.02-1.42) |
| **19** | 1.00 (0.99-1.02) | 0.89 (0.88-0.90) | 0.90 (0.89-0.91) | 0.51 (0.48-0.54) | 0.75 (0.72-0.78) | 0.77 (0.74-0.81) | 0.53 (0.51-0.56) | 0.69 (0.65-0.73) | 0.71 (0.68-0.75) | 0.95 (0.92-0.98) | 1.13 (1.03-1.23) | 1.13 (1.04-1.23) | 1.04 (1.00-1.07) | 1.08 (1.00-1.17) | 1.08 (1.00-1.16) |
| **22** | reference | reference | reference | reference | reference | reference | reference | reference | reference | reference | reference | reference | reference | reference | reference |
| **25** | 1.00 (0.99-1.01) | 1.11 (1.10-1.12) | 1.10 (1.09-1.11) | 1.85 (1.76-1.95) | 1.30 (1.26-1.34) | 1.27 (1.22-1.31) | 1.77 (1.69-1.85) | 1.41 (1.35-1.48) | 1.37 (1.31-1.43) | 1.05 (1.02-1.09) | 0.91 (0.84-0.98) | 0.90 (0.84-0.97) | 0.97 (0.93-1.00) | 0.92 (0.86-1.00) | 0.97 (0.91-1.03) |
| **28** | 1.02 (1.00-1.04) | 1.20 (1.18-1.22) | 1.17 (1.15-1.19) | 2.79 (2.58-3.01) | 1.64 (1.55-1.74) | 1.56 (1.47-1.66) | 2.60 (2.42-2.79) | 1.83 (1.70-1.97) | 1.74 (1.61-1.87) | 1.11 (1.05-1.18) | 0.88 (0.78-0.99) | 0.88 (0.78-0.99) | 0.93 (0.87-0.99) | 0.89 (0.78-1.02) | 0.97 (0.88-1.08) |
| **31** | 1.06 (1.04-1.08) | 1.24 (1.22-1.27) | 1.22 (1.19-1.24) | 3.34 (3.08-3.63) | 2.01 (1.87-2.17) | 1.88 (1.75-2.03) | 3.06 (2.83-3.30) | 2.14 (1.98-2.31) | 2.01 (1.86-2.18) | 1.17 (1.07-1.29) | 0.93 (0.82-1.06) | 0.93 (0.82-1.05) | 0.90 (0.82-0.99) | 0.94 (0.80-1.10) | 1.02 (0.89-1.17) |
| **34** | 1.11 (1.09-1.13) | 1.27 (1.25-1.30) | 1.24 (1.22-1.26) | 3.58 (3.30-3.88) | 2.40 (2.21-2.60) | 2.22 (2.04-2.41) | 3.19 (2.96-3.44) | 2.37 (2.19-2.56) | 2.22 (2.06-2.40) | 1.24 (1.09-1.40) | 1.03 (0.90-1.18) | 1.02 (0.89-1.17) | 0.87 (0.76-0.99) | 1.08 (0.92-1.26) | 1.11 (0.95-1.31) |
| **37** | 1.16 (1.13-1.19) | 1.29 (1.26-1.33) | 1.26 (1.23-1.29) | 3.77 (3.46-4.11) | 2.78 (2.54-3.03) | 2.54 (2.33-2.78) | 3.26 (3.01-3.53) | 2.59 (2.39-2.81) | 2.43 (2.24-2.64) | 1.30 (1.12-1.52) | 1.15 (0.98-1.36) | 1.14 (0.97-1.34) | 0.84 (0.71-0.98) | 1.28 (1.09-1.51) | 1.26 (1.06-1.51) |
| **40** | 1.21 (1.18-1.25) | 1.32 (1.28-1.36) | 1.28 (1.25-1.32) | 3.97 (3.61-4.37) | 3.12 (2.84-3.44) | 2.85 (2.58-3.13) | 3.32 (3.03-3.63) | 2.83 (2.59-3.10) | 2.66 (2.43-2.91) | 1.38 (1.14-1.66) | 1.29 (1.05-1.58) | 1.27 (1.03-1.56) | 0.81 (0.67-0.98) | 1.52 (1.26-1.84) | 1.49 (1.23-1.81) |
| **43** | 1.27 (1.23-1.32) | 1.35 (1.30-1.40) | 1.31 (1.26-1.36) | 4.18 (3.75-4.67) | 3.41 (3.04-3.82) | 3.11 (2.77-3.49) | 3.38 (3.05-3.75) | 3.10 (2.80-3.44) | 2.91 (2.62-3.23) | 1.45 (1.17-1.80) | 1.44 (1.11-1.85) | 1.42 (1.10-1.83) | 0.78 (0.62-0.98) | 1.81 (1.45-2.27) | 1.83 (1.47-2.29) |
| **47** | 1.35 (1.29-1.42) | 1.38 (1.32-1.45) | 1.34 (1.28-1.40) | 4.48 (3.92-5.12) | 3.66 (3.12-4.31) | 3.37 (2.87-3.96) | 3.47 (3.06-3.94) | 3.50 (3.08-3.97) | 3.27 (2.88-3.72) | 1.56 (1.21-2.01) | 1.67 (1.20-2.30) | 1.64 (1.18-2.27) | 0.74 (0.57-0.97) | 2.29 (1.72-3.04) | 2.56 (1.92-3.41) |
| **50** | 1.42 (1.35-1.50) | 1.41 (1.33-1.48) | 1.36 (1.29-1.44) | 4.72 (4.05-5.50) | 3.74 (3.01-4.64) | 3.48 (2.81-4.31) | 3.54 (3.06-4.10) | 3.83 (3.31-4.43) | 3.58 (3.09-4.15) | 1.64 (1.23-2.19) | 1.86 (1.27-2.72) | 1.82 (1.24-2.68) | 0.72 (0.53-0.97) | 2.72 (1.95-3.80) | 3.45 (2.41-4.96) |

Notes: the fully adjusted models are adjusted for age, sex, smoking status and the MEDEA deprivation index.
Abbreviations: BMI: Body mass index; CI: Confidence interval; COVID-19: Coronavirus Disease 2019.

**Table S4.** Hazards ratios of COVID-19 outcomes related to body mass index, with 95% CIs, stratified by age and sex

A. Age

| | **From general population** | | | | | | **From diagnosed with COVID-19** | | | | | | **From hospitalised with COVID-19** | | |
|---|---|---|---|---|---|---|---|---|---|---|---|---|---|---|---|
| | **to diagnosed with COVID-19** | | | **to hospitalised with COVID-19** | | | **to hospitalised with COVID-19** | | | **to death** | | | **to death** | | |
| **BMI values (kg/m$^2$)** | **Aged 18 to 59** | **Aged 60 to 79** | **Aged 80 or older** | **Aged 18 to 59** | **Aged 60 to 79** | **Aged 80 or older** | **Aged 18 to 59** | **Aged 60 to 79** | **Aged 80 or older** | **Aged 18 to 59** | **Aged 60 to 79** | **Aged 80 or older** | **Aged 18 to 59** | **Aged 60 to 79** | **Aged 80 or older** |
| **16** | 0.80<br>(0.77-0.82) | 0.97<br>(0.95-0.99) | 1.23<br>(1.11-1.36) | 0.39<br>(0.33-0.47) | 0.79<br>(0.76-0.83) | 0.84<br>(0.77-0.90) | 0.44<br>(0.38-0.52) | 0.81<br>(0.77-0.85) | 0.91<br>(0.83-0.99) | 6.37<br>(2.22-18.23) | 1.95<br>(1.39-2.73) | 1.05<br>(0.97-1.14) | 0.64<br>(0.52-0.80) | 1.27<br>(0.99-1.63) | 0.98<br>(0.88-1.09) |
| **19** | 0.89<br>(0.88-0.91) | 0.98<br>(0.97-1.00) | 1.11<br>(1.05-1.17) | 0.64<br>(0.60-0.69) | 0.89<br>(0.87-0.91) | 0.91<br>(0.88-0.95) | 0.67<br>(0.62-0.72) | 0.90<br>(0.88-0.92) | 0.95<br>(0.91-1.00) | 2.52<br>(1.49-4.26) | 1.40<br>(1.18-1.65) | 1.03<br>(0.98-1.07) | 0.80<br>(0.72-0.89) | 1.10<br>(0.98-1.23) | 0.99<br>(0.94-1.04) |
| **22** | reference | reference | reference | reference | reference | reference | reference | reference | reference | reference | reference | reference | reference | reference | reference |
| **25** | 1.10<br>(1.09-1.12) | 1.02<br>(1.00-1.03) | 0.91<br>(0.87-0.96) | 1.49<br>(1.40-1.58) | 1.12<br>(1.10-1.15) | 1.09<br>(1.05-1.14) | 1.45<br>(1.36-1.55) | 1.11<br>(1.08-1.14) | 1.05<br>(1.00-1.10) | 0.47<br>(0.30-0.74) | 0.72<br>(0.61-0.85) | 0.97<br>(0.94-1.02) | 1.25<br>(1.12-1.39) | 0.95<br>(0.87-1.04) | 1.01<br>(0.96-1.07) |
| **28** | 1.18<br>(1.16-1.20) | 1.03<br>(1.01-1.06) | 0.90<br>(0.83-0.97) | 2.12<br>(1.91-2.35) | 1.26<br>(1.21-1.32) | 1.20<br>(1.11-1.29) | 1.91<br>(1.72-2.12) | 1.24<br>(1.18-1.30) | 1.10<br>(1.01-1.21) | 0.36<br>(0.18-0.73) | 0.59<br>(0.44-0.78) | 0.95<br>(0.87-1.03) | 1.55<br>(1.25-1.92) | 0.95<br>(0.81-1.12) | 1.02<br>(0.92-1.14) |
| **31** | 1.23<br>(1.20-1.25) | 1.05<br>(1.01-1.08) | 0.98<br>(0.91-1.06) | 2.88<br>(2.52-3.29) | 1.42<br>(1.33-1.52) | 1.31<br>(1.17-1.47) | 2.26<br>(2.02-2.53) | 1.37<br>(1.27-1.48) | 1.16<br>(1.01-1.33) | 0.46<br>(0.22-0.98) | 0.61<br>(0.44-0.84) | 0.93<br>(0.82-1.05) | 1.93<br>(1.40-2.66) | 1.00<br>(0.81-1.23) | 1.03<br>(0.88-1.22) |
| **34** | 1.26<br>(1.23-1.29) | 1.07<br>(1.02-1.11) | 1.13<br>(1.04-1.23) | 3.74<br>(3.21-4.35) | 1.60<br>(1.46-1.75) | 1.43<br>(1.23-1.67) | 2.55<br>(2.28-2.85) | 1.53<br>(1.38-1.68) | 1.22<br>(1.01-1.46) | 0.77<br>(0.37-1.58) | 0.74<br>(0.54-1.02) | 0.90<br>(0.77-1.07) | 2.41<br>(1.57-3.69) | 1.10<br>(0.86-1.40) | 1.05<br>(0.84-1.30) |
| **37** | 1.29<br>(1.26-1.32) | 1.08<br>(1.02-1.15) | 1.31<br>(1.18-1.46) | 4.64<br>(3.95-5.45) | 1.80<br>(1.60-2.02) | 1.56<br>(1.29-1.89) | 2.84<br>(2.54-3.18) | 1.70<br>(1.50-1.92) | 1.28<br>(1.02-1.61) | 1.33<br>(0.65-2.75) | 0.96<br>(0.68-1.35) | 0.88<br>(0.72-1.08) | 3.00<br>(1.76-5.12) | 1.27<br>(0.97-1.65) | 1.06<br>(0.81-1.39) |
| **40** | 1.32<br>(1.28-1.37) | 1.10<br>(1.03-1.18) | 1.52<br>(1.33-1.74) | 5.50<br>(4.64-6.52) | 2.02<br>(1.76-2.32) | 1.71<br>(1.36-2.15) | 3.17<br>(2.80-3.59) | 1.88<br>(1.62-2.19) | 1.34<br>(1.02-1.77) | 2.32<br>(1.08-4.97) | 1.23<br>(0.82-1.85) | 0.86<br>(0.67-1.10) | 3.74<br>(1.97-7.09) | 1.53<br>(1.15-2.04) | 1.07<br>(0.77-1.48) |
| **43** | 1.35<br>(1.30-1.41) | 1.12<br>(1.03-1.21) | 1.76<br>(1.49-2.08) | 6.24<br>(5.18-7.51) | 2.27<br>(1.93-2.67) | 1.87<br>(1.43-2.44) | 3.54<br>(3.07-4.08) | 2.09<br>(1.76-2.49) | 1.41<br>(1.02-1.94) | 4.04<br>(1.75-9.33) | 1.58<br>(0.96-2.59) | 0.84<br>(0.63-1.12) | 4.66<br>(2.20-9.83) | 1.95<br>(1.42-2.66) | 1.08<br>(0.74-1.58) |
| **47** | 1.40<br>(1.33-1.47) | 1.14<br>(1.04-1.25) | 2.15<br>(1.74-2.66) | 6.87<br>(5.43-8.70) | 2.65<br>(2.19-3.22) | 2.11<br>(1.53-2.90) | 4.09<br>(3.45-4.85) | 2.41<br>(1.96-2.96) | 1.51<br>(1.03-2.20) | 8.46<br>(3.18-22.52) | 2.20<br>(1.17-4.13) | 0.81<br>(0.57-1.14) | 6.24<br>(2.56-15.2) | 2.88<br>(1.95-4.24) | 1.10<br>(0.70-1.73) |
| **50** | 1.43<br>(1.35-1.52) | 1.16<br>(1.04-1.29) | 2.49<br>(1.95-3.19) | 7.01<br>(5.18-9.48) | 2.98<br>(2.41-3.70) | 2.30<br>(1.61-3.29) | 4.56<br>(3.76-5.55) | 2.68<br>(2.13-3.38) | 1.58<br>(1.03-2.42) | 14.72<br>(4.86-44.52) | 2.83<br>(1.35-5.91) | 0.79<br>(0.54-1.16) | 7.77<br>(2.87-21.06) | 4.08<br>(2.53-6.60) | 1.11<br>(0.67-1.85) |

B. Sex

| | From general population | | | | From diagnosed with COVID-19 | | | | From hospitalised with COVID-19 | |
|---|---|---|---|---|---|---|---|---|---|---|
| | to diagnosed with COVID-19 | | to hospitalised with COVID-19 | | to hospitalised with COVID-19 | | to death | | to death | |
| **BMI values (kg/m$^2$)** | **Females** | **Males** | **Females** | **Males** | **Females** | **Males** | **Females** | **Males** | **Females** | **Males** |
| **16** | 0.84 (0.82-0.87) | 0.74 (0.70-0.77) | 0.51 (0.45-0.58) | 0.77 (0.73-0.8) | 0.49 (0.43-0.57) | 0.54 (0.46-0.62) | 0.98 (0.90-1.07) | 0.93 (0.82-1.06) | 1.25 (1.01-1.56) | 0.93 (0.84-1.02) |
| **19** | 0.92 (0.90-0.93) | 0.86 (0.84-0.88) | 0.73 (0.68-0.77) | 0.88 (0.86-0.9) | 0.7 (0.66-0.75) | 0.73 (0.68-0.79) | 0.99 (0.95-1.03) | 0.97 (0.91-1.03) | 1.12 (1.00-1.25) | 0.96 (0.92-1.01) |
| **22** | reference | reference | reference | reference | reference | reference | reference | reference | reference | reference |
| **25** | 1.08 (1.07-1.10) | 1.16 (1.13-1.18) | 1.34 (1.28-1.40) | 1.14 (1.12-1.17) | 1.38 (1.3-1.47) | 1.35 (1.26-1.45) | 1.01 (0.97-1.05) | 1.03 (0.97-1.10) | 0.90 (0.81-1.00) | 1.04 (0.99-1.09) |
| **28** | 1.14 (1.12-1.17) | 1.28 (1.23-1.32) | 1.74 (1.60-1.88) | 1.3 (1.24-1.37) | 1.74 (1.59-1.92) | 1.73 (1.54-1.93) | 1.02 (0.93-1.11) | 1.07 (0.94-1.22) | 0.85 (0.70-1.03) | 1.08 (0.98-1.19) |
| **31** | 1.19 (1.16-1.21) | 1.33 (1.28-1.37) | 2.19 (1.97-2.43) | 1.49 (1.39-1.59) | 2.01 (1.82-2.23) | 2.01 (1.78-2.27) | 1.03 (0.90-1.17) | 1.11 (0.92-1.34) | 0.90 (0.71-1.14) | 1.12 (0.96-1.30) |
| **34** | 1.22 (1.19-1.24) | 1.34 (1.29-1.39) | 2.67 (2.38-3.01) | 1.7 (1.55-1.86) | 2.21 (1.99-2.45) | 2.25 (2.00-2.52) | 1.04 (0.87-1.23) | 1.15 (0.89-1.48) | 1.07 (0.84-1.36) | 1.16 (0.95-1.42) |
| **37** | 1.24 (1.21-1.28) | 1.35 (1.29-1.41) | 3.17 (2.80-3.60) | 1.94 (1.73-2.18) | 2.39 (2.15-2.66) | 2.49 (2.20-2.81) | 1.04 (0.84-1.29) | 1.19 (0.86-1.63) | 1.35 (1.06-1.72) | 1.21 (0.94-1.55) |
| **40** | 1.27 (1.23-1.32) | 1.36 (1.29-1.43) | 3.65 (3.19-4.17) | 2.21 (1.93-2.54) | 2.59 (2.3-2.92) | 2.76 (2.40-3.17) | 1.05 (0.82-1.36) | 1.23 (0.84-1.80) | 1.74 (1.34-2.26) | 1.26 (0.93-1.69) |
| **43** | 1.30 (1.24-1.36) | 1.37 (1.28-1.46) | 4.08 (3.51-4.74) | 2.53 (2.15-2.97) | 2.81 (2.45-3.23) | 3.06 (2.60-3.59) | 1.06 (0.79-1.43) | 1.27 (0.82-1.98) | 2.23 (1.66-3.00) | 1.30 (0.92-1.85) |
| **47** | 1.34 (1.27-1.42) | 1.38 (1.27-1.50) | 4.51 (3.69-5.50) | 3.02 (2.49-3.66) | 3.13 (2.64-3.7) | 3.51 (2.88-4.27) | 1.08 (0.75-1.54) | 1.33 (0.78-2.25) | 3.11 (2.16-4.48) | 1.37 (0.90-2.08) |
| **50** | 1.37 (1.28-1.46) | 1.39 (1.26-1.53) | 4.69 (3.62-6.08) | 3.44 (2.77-4.27) | 3.39 (2.79-4.11) | 3.89 (3.10-4.88) | 1.09 (0.73-1.62) | 1.38 (0.76-2.48) | 4.00 (2.62-6.10) | 1.42 (0.89-2.27) |

Notes: the models are adjusted for age, sex, smoking status and the MEDEA deprivation index.
Abbreviations: BMI: Body mass index; CI: Confidence interval; COVID-19: Coronavirus Disease 2019.

**Figure S4**. Association between body mass index and the risk of COVID-19 diagnosis, allowing for non-linear effects, with 95% CIs, stratified by calendar month

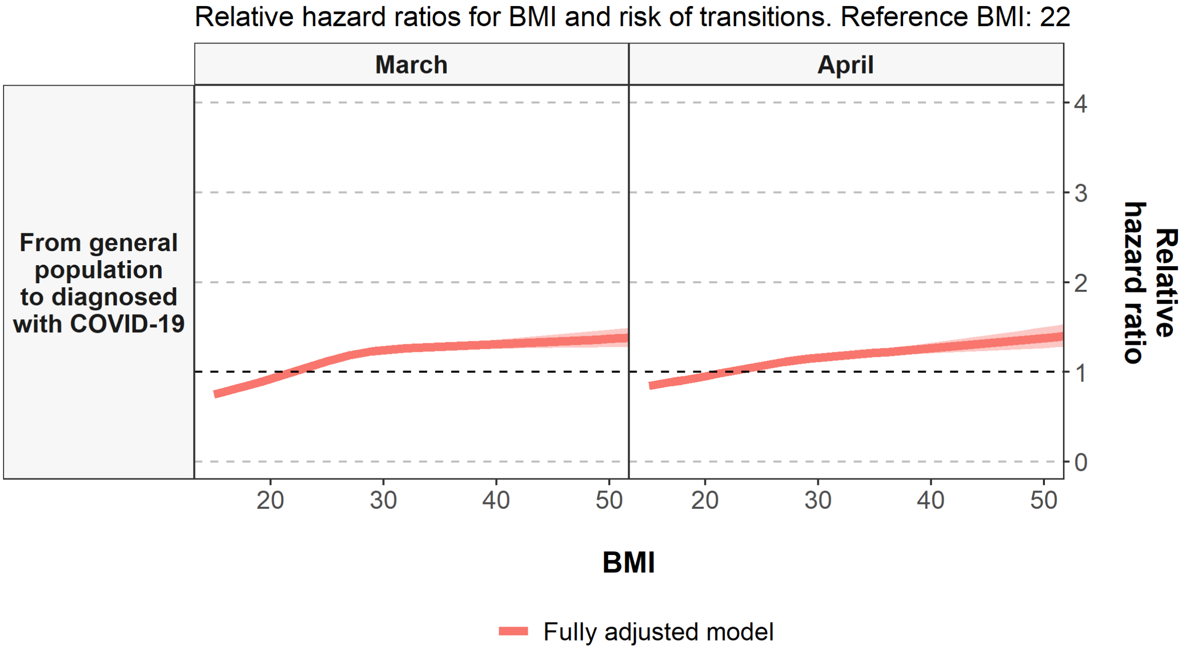


Notes: Models are adjusted for age, sex, smoking status and the MEDEA deprivation index. These models were stratified by calendar month to account for the violation of the assumption of proportionality for age in the first transition.

Abbreviations: BMI: Body mass index; CI: Confidence interval; COVID-19: Coronavirus Disease 2019.

**Table S5.** Hazard Ratios of the risk of COVID-19 diagnosis related to body mass index, allowing for non-linear effects, with 95% CIs, stratified by calendar month

| | From general population to diagnosed with COVID-19 | | |
|---|---|---|---|
| **BMI values ($kg/m^2$)** | **Overall** | **March** | **April** |
| **16** | 0.81 (0.79-0.84) | 0.78 (0.75-0.81) | 0.86 (0.83-0.90) |
| **19** | 0.90 (0.89-0.91) | 0.88 (0.87-0.90) | 0.93 (0.91-0.95) |
| **22** | reference | reference | reference |
| **25** | 1.10 (1.09-1.11) | 1.12 (1.10-1.14) | 1.07 (1.05-1.09) |
| **28** | 1.17 (1.15-1.19) | 1.21 (1.18-1.24) | 1.13 (1.10-1.16) |
| **31** | 1.22 (1.19-1.24) | 1.25 (1.22-1.28) | 1.17 (1.14-1.20) |
| **34** | 1.24 (1.22-1.26) | 1.27 (1.24-1.31) | 1.20 (1.16-1.24) |
| **37** | 1.26 (1.23-1.29) | 1.29 (1.25-1.33) | 1.23 (1.19-1.28) |
| **40** | 1.28 (1.25-1.32) | 1.31 (1.26-1.36) | 1.26 (1.21-1.32) |
| **43** | 1.31 (1.26-1.36) | 1.33 (1.26-1.39) | 1.30 (1.23-1.37) |
| **47** | 1.34 (1.28-1.40) | 1.35 (1.27-1.43) | 1.34 (1.25-1.44) |
| **50** | 1.36 (1.29-1.44) | 1.37 (1.27-1.47) | 1.38 (1.27-1.50) |

Notes: Models are adjusted for age, sex, smoking status and the MEDEA deprivation index. These models were stratified by calendar month to account for the violation of the assumption of proportionality for age in the first transition.
Abbreviations: BMI: Body mass index; CI: Confidence interval; COVID-19: Coronavirus Disease 2019.

**Figure S5.** Hazard Ratios of COVID-19 outcomes related to body mass index in categories, with 95% CIs

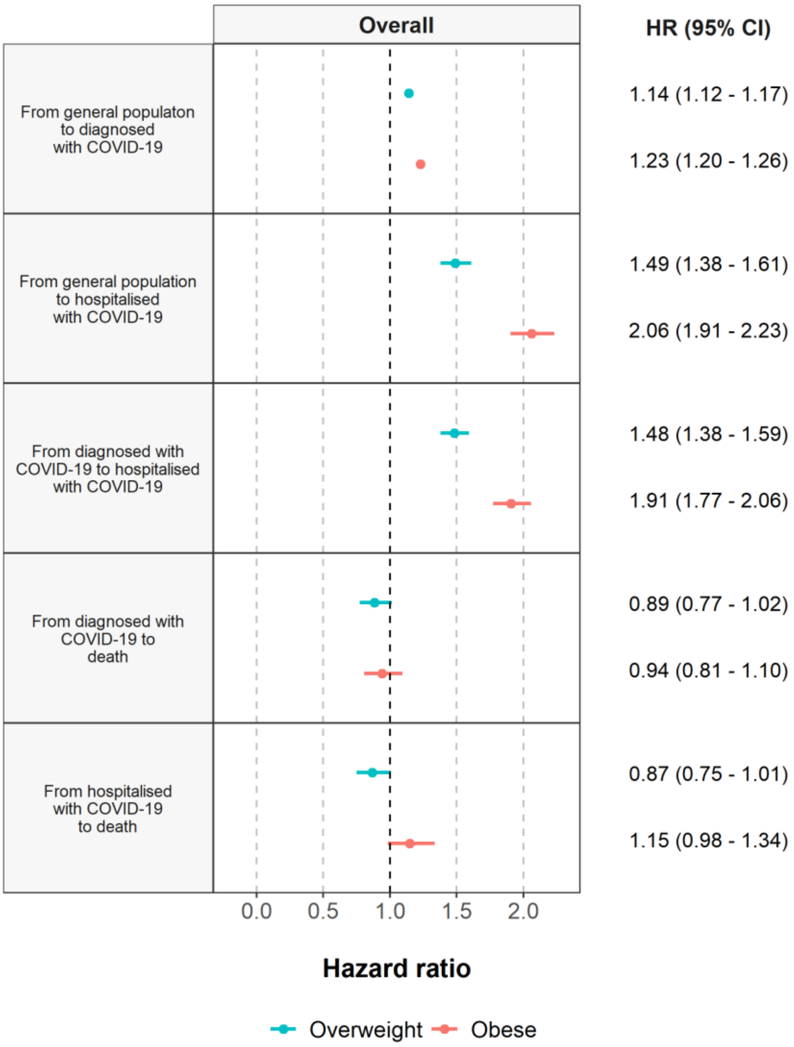


Notes: Hazard ratios are relative to normal or underweight (BMI <18.5 kg/m$^2$ and between ≥18.5 and <25 kg/m$^2$). The overweight category includes BMIs between ≥25 and <30 kg/m$^2$ and for the obesity category BMIs ≥30 kg/m$^2$. Models are adjusted for age, sex, smoking status and the MEDEA deprivation index.
Abbreviations: BMI: Body mass index; CI: Confidence interval; COVID-19: Coronavirus Disease 2019; HR: Hazard ratio.

**Figure S6.** Association between body mass index and the risk of COVID-19 outcomes, allowing for non-linear effects, with 95% CIs, after multiple imputation of missing data on BMI, smoking status, and/or the MEDEA deprivation index

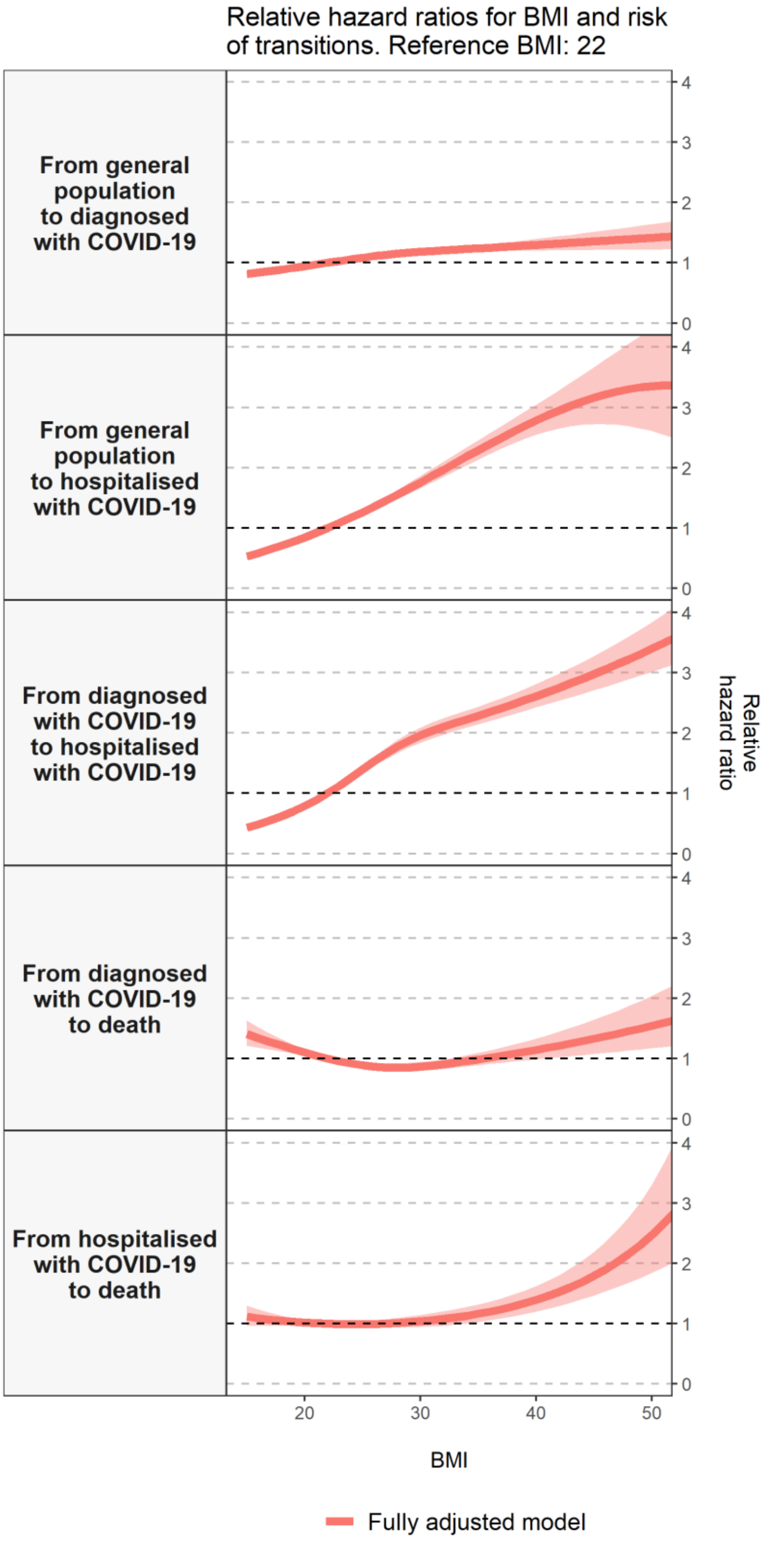


Notes: Models are adjusted for age, sex, smoking status and the MEDEA deprivation index. For the multiple imputations we used predictive mean matching, with 5 imputations drawn.
Abbreviations: BMI: Body mass index; CI: Confidence interval; COVID-19: Coronavirus Disease 2019; MEDEA: "Mortalidad en áreas pequeñas Españolas y Desigualdades Socioeconómicas y Ambientales".

**Table S6.** Hazards ratios of COVID-19 outcomes related to body mass index, with 95% CIs, after multiple imputation of missing data on BMI, smoking status, and/or the MEDEA deprivation index

| | **From General Population** | | **From diagnosed with COVID-19** | | **From hospitalised with COVID-19** |
|---|---|---|---|---|---|
| **BMI values (kg/m$^2$)** | **To diagnosed with COVID-19** | **To hospitalised with COVID-19** | **To hospitalised with COVID-19** | **To death** | **To death** |
| **16** | 0.84 (0.79-0.88) | 0.58 (0.53-0.64) | 0.49 (0.44-0.53) | 1.34 (1.18-1.52) | 1.09 (0.96-1.24) |
| **19** | 0.91 (0.89-0.94) | 0.77 (0.74-0.81) | 0.70 (0.67-0.73) | 1.16 (1.09-1.23) | 1.03 (0.97-1.09) |
| **22** | reference | reference | reference | reference | reference |
| **25** | 1.09 (1.06-1.11) | 1.26 (1.22-1.30) | 1.39 (1.34-1.45) | 0.88 (0.84-0.93) | 0.99 (0.95-1.04) |
| **28** | 1.15 (1.12-1.18) | 1.55 (1.48-1.63) | 1.77 (1.67-1.88) | 0.85 (0.78-0.92) | 1.01 (0.94-1.10) |
| **31** | 1.20 (1.16-1.23) | 1.86 (1.75-1.98) | 2.04 (1.91-2.17) | 0.88 (0.81-0.96) | 1.06 (0.95-1.17) |
| **34** | 1.23 (1.19-1.27) | 2.18 (2.04-2.34) | 2.23 (2.09-2.37) | 0.96 (0.87-1.05) | 1.13 (1.00-1.28) |
| **37** | 1.26 (1.20-1.33) | 2.50 (2.32-2.69) | 2.41 (2.26-2.57) | 1.05 (0.93-1.18) | 1.24 (1.08-1.42) |
| **40** | 1.30 (1.20-1.39) | 2.78 (2.55-3.04) | 2.61 (2.42-2.81) | 1.14 (0.98-1.33) | 1.40 (1.20-1.62) |
| **43** | 1.33 (1.21-1.46) | 3.03 (2.69-3.41) | 2.82 (2.59-3.08) | 1.25 (1.04-1.51) | 1.61 (1.35-1.92) |
| **47** | 1.38 (1.22-1.56) | 3.26 (2.71-3.93) | 3.14 (2.83-3.49) | 1.41 (1.11-1.79) | 2.02 (1.60-2.55) |
| **50** | 1.41 (1.22-1.64) | 3.35 (2.61-4.31) | 3.40 (3.01-3.84) | 1.54 (1.17-2.04) | 2.47 (1.84-3.31) |

Notes: Models are adjusted for age, sex, smoking status and the MEDEA deprivation index. For the multiple imputations we used predictive mean matching, with 5 imputations drawn.

Abbreviations: BMI: Body mass index; CI: Confidence interval; COVID-19: Coronavirus Disease 2019; MEDEA: “Mortalidad en áreas pequeñas Españolas y Desigualdades Socioeconómicas y Ambientales”.

**Figure S7.** Association between body mass index and the risk of COVID-19 outcomes, allowing for non-linear effects, with 95% CIs among people with a BMI measurement not older than five years prior to the index date

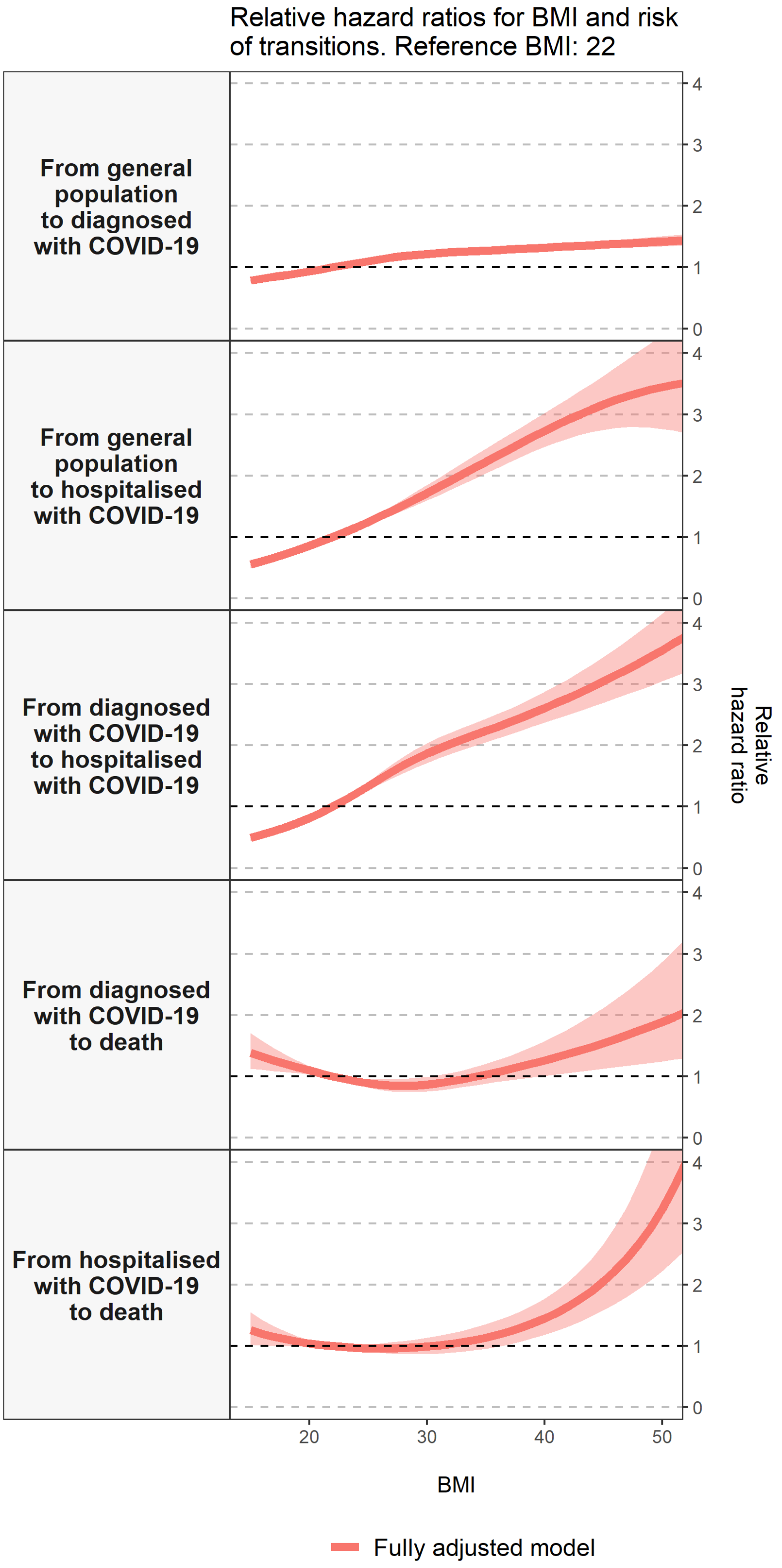


Notes: Models are adjusted for age, sex, smoking status and the MEDEA deprivation index
Abbreviations: BMI: Body mass index; CI: Confidence interval; COVID-19: Coronavirus Disease 2019.

**Table S7.** Hazards ratios of COVID-19 outcomes related to body mass index, with 95% CIs among people with a BMI measurement not older than five years prior to the index date

| | From General Population | | From diagnosed with COVID-19 | | From hospitalised with COVID-19 |
|---|---|---|---|---|---|
| BMI values (kg/m$^2$) | To diagnosed with COVID-19 | To hospitalised with COVID-19 | To hospitalised with COVID-19 | To death | To death |
| **16** | 0.81 (0.79-0.84) | 0.61 (0.55-0.67) | 0.55 (0.49-0.61) | 1.32 (1.11-1.58) | 1.20 (1.01-1.43) |
| **19** | 0.90 (0.89-0.92) | 0.79 (0.75-0.82) | 0.74 (0.70-0.78) | 1.15 (1.05-1.26) | 1.08 (0.99-1.16) |
| **22** | reference | reference | reference | reference | reference |
| **25** | 1.10 (1.09-1.11) | 1.25 (1.20-1.29) | 1.33 (1.27-1.40) | 0.89 (0.82-0.96) | 0.97 (0.91-1.03) |
| **28** | 1.18 (1.16-1.21) | 1.52 (1.43-1.62) | 1.68 (1.55-1.81) | 0.85 (0.75-0.96) | 0.97 (0.87-1.08) |
| **31** | 1.23 (1.21-1.26) | 1.82 (1.68-1.96) | 1.95 (1.79-2.13) | 0.89 (0.77-1.02) | 1.01 (0.88-1.17) |
| **34** | 1.26 (1.24-1.29) | 2.12 (1.95-2.32) | 2.17 (1.99-2.36) | 0.99 (0.85-1.14) | 1.10 (0.93-1.30) |
| **37** | 1.29 (1.26-1.33) | 2.43 (2.22-2.67) | 2.38 (2.18-2.59) | 1.12 (0.93-1.33) | 1.24 (1.03-1.49) |
| **40** | 1.32 (1.28-1.36) | 2.73 (2.47-3.02) | 2.61 (2.37-2.87) | 1.26 (1.01-1.58) | 1.45 (1.18-1.78) |
| **43** | 1.35 (1.30-1.40) | 3.00 (2.66-3.38) | 2.86 (2.56-3.20) | 1.42 (1.08-1.88) | 1.76 (1.40-2.22) |
| **47** | 1.39 (1.32-1.46) | 3.29 (2.79-3.89) | 3.24 (2.83-3.71) | 1.67 (1.17-2.39) | 2.43 (1.80-3.28) |
| **50** | 1.42 (1.34-1.50) | 3.45 (2.76-4.30) | 3.55 (3.05-4.15) | 1.89 (1.25-2.87) | 3.24 (2.22-4.71) |

Notes: Models are adjusted for age, sex, smoking status and the MEDEA deprivation index
Abbreviations: BMI: Body mass index; CI: Confidence interval; COVID-19: Coronavirus Disease 2019.
